# Altered T1w/T2w-FLAIR Ratio in White Matter Hyperintensities as an Indicator of Structural Integrity Loss: Association with Alzheimer’s Disease and Vascular Dementia

**DOI:** 10.64898/2026.07.28.26359139

**Authors:** Rushil Srirambhatla, Jacques-Yves Campion, Thomas Desmidt, Yiyan Pan, Carmen Andreescu, Pamela C.L. Ferreira, Guilherme Povala, Bruna Bellaver, João Pedro Ferrari-Souza, Douglas T. Leffa, Firoza Z. Lussier, Marina Scop Medeiros, Emma Ruppert, Francieli Rohden, Chang Hyung Hong, Hyun Woong Roh, Bumhee Park, Jin Wook Choi, Sang Won Seo, Seong Hye Choi, So Young Moon, Eun-Joo Kim, Byeong C. Kim, Young-Sil An, Yong Hyuk Cho, Sunhwa Hong, Thomas K. Karikari, Tharick A. Pascoal, Sang Joon Son, Helmet T. Karim

## Abstract

**Background:** White matter hyperintensities (WMH) are prevalent in dementia, but lesion volume does not capture their microstructural heterogeneity. The T1-weighted to fluid-attenuated inversion recovery (T1w/T2w-FLAIR) ratio is sensitive to myelin, gliosis, and tissue water. We tested whether lesion-specific T1w/T2w-FLAIR ratio, referenced to each participant’s normal-appearing white matter (NAWM), differs by diagnosis and reflects distinct amyloid and vascular mechanisms in Alzheimer’s disease (AD) and vascular dementia (VD).

**Methods:** We analyzed 576 participants from the multicentre BICWALZS cohort (seven South Korean sites), spanning subjective cognitive impairment (SCI, n=71), mild cognitive impairment (n=270), AD (n=125) and VD (n=88). WMH T1w/T2w-FLAIR ratio was regressed on NAWM ratio, yielding standardized residuals as the outcome. Regression and mediation models tested diagnosis, plasma biomarkers, APOE genotype, amyloid PET and vascular risk burden, adjusting for age, sex, education and site. We conducted regression and mediation analyses after multiple imputation for missing variables. We adjusted for hierarchical models using Bonferroni correction. We tested for insensitivity to site effects by applying ComBat harmonization.

**Results:** Older age, AD, VD, and high vascular risk burden were associated with higher residualized T1w/T2w-FLAIR ratios relative to SCI. Lower plasma amyloid-beta 42 (greater amyloid burden) was associated with lower T1w/T2w-FLAIR ratios. Greater vascular burden was associated with greater T1w/T2w-FLAIR ratios, which partially mediated the VD association with T1w/T2w-FLAIR. Lower amyloid-beta 42 (greater amyloid) was associated with lower T1w/T2w-FLAIR, which partially mediated the effect between AD and T1w/T2w-FLAIR ratio. Findings were robust to harmonization.

**Conclusions:** Residualized WMH T1w/T2w-FLAIR ratio captures lesion-specific microstructural variation missed by volumetric measures, consistent with vascular-gliotic injury in VD and coexisting amyloid-linked demyelination in AD. Limitations include the cross-sectional design, no cognitively normal comparison group, and a predominantly Korean sample.

## 1. Background

White matter hyperintensities (WMH) on T2-weighted magnetic resonance imaging (MRI) are a highly prevalent marker of small vessel disease in older adults. These lesions, present in over 90% of adults aged 65 and older (Bahrani et al., 2025), predict increased risk of stroke, cognitive decline, and both vascular dementia (VD) and Alzheimer-type dementia (AD) (Launer et al. 2005; De Leeuw et al. 2001; Wardlaw et al. 2013). Conventional WMH quantification methods focus predominantly on lesion burden, the total volume of affected tissue, while providing limited insight into the microstructural integrity of tissue within WMH (Wardlaw et al. 2013). This gap is particularly critical in mixed-pathology dementia cohorts, where the relative contributions of vascular injury (including microvascular ischemic events leading to myelin damage) and Alzheimer-related white matter changes (such as white matter rarefaction or reduced myelin density associated with AD histopathology) are not well characterized. Whether vascular and Alzheimer-related injury leave distinguishable signatures within the lesions themselves, rather than differing only in lesion burden, remains unknown.

The T1-weighted to T2-weighted (T1w/T2w) ratio has emerged as a composite marker sensitive to myelin, water content, gliosis, and tissue damage, with its biological interpretation depending on disease context. Originally developed to map cortical myelin content in healthy individuals, the T1w/T2w ratio captures relative contrast related to myelin concentration, axonal integrity, and tissue organization (Glasser and Van Essen 2011). WMH reflect a diverse spectrum of pathological changes that collectively influence T1 and T2 relaxation times and, by extension, the T1w/T2w ratio. At the tissue level, these include axonal loss, reactive gliosis, oligodendrocyte degeneration, neuroinflammation, and microglial activation (Hase et al., 2018; Erten-Lyons et al., 2013; Haller et al., 2013; Iordanishvili et al., 2019). At the vascular and fluid level, they include arteriolosclerosis, venular collagenosis, enlarged perivascular spaces, and increased interstitial water content from blood-brain barrier (BBB) disruption (Roseborough et al., 2022; Solé-Guardia et al., 2024; Wardlaw et al., 2019; Joutel & Chabriat, 2017; Silbert et al., 2024). In AD, WMH may also be associated with cerebral amyloid angiopathy and neurodegenerative changes, where vascular and AD-related mechanisms overlap (Erten-Lyons et al., 2013; Shirzadi et al., 2023).

Because the T1w/T2w ratio is sensitive to variations in myelin content, tissue water, and cellular composition, it is well-positioned to capture this pathological heterogeneity – potentially differentiating WMH with predominantly vascular injury from those with superimposed neurodegenerative changes.

In multiple sclerosis (MS), the T1w/T2w ratio has been validated as sensitive to microstructural injury, with reduced ratios tracking demyelination and tissue disruption within lesions (Pareto et al., 2020; Cooper et al., 2019; Cappelle et al., 2022), and longitudinal changes correlating with disease activity and disability (Boaventura et al., 2022). This approach rests on histological work establishing that MRI signal intensity in high-resolution structural images reflects underlying cortical myeloarchitecture and cytoarchitecture (Eickhoff et al., 2005). Postmortem studies combining MRI with histopathology have since linked the T1w/T2w ratio directly to demyelination, with significantly reduced ratios in histologically confirmed demyelinated cortex relative to myelinated cortex in MS (Nakamura et al., 2017). A further postmortem study anchored cortical T1w/T2w values to underlying tissue composition, although dendrite density rather than myelin content emerged as the dominant contributor in that work (Righart et al., 2017). Histological specificity in white matter specifically remains debated, with some studies reporting weaker or discrepant correspondence between T1w/T2w and myelin measures than quantitative relaxometry achieves. We therefore interpret the ratio throughout as a composite index of myelin, gliosis, and tissue water rather than a myelin-specific measure. Translation of this technique to aging cohorts with WMH and mixed vascular–neurodegenerative pathology has been limited, and it remains unclear whether T1w/T2w-FLAIR–indexed microstructural differences within WMH relate specifically to molecular markers of disease.

Although WMH appear similar on imaging across dementia subtypes, they arise from distinct pathological processes. In VD, lesions follow a relatively stereotyped sequence of chronic hypoperfusion, ischemic demyelination, axonal loss, oligodendrocyte depletion, reactive gliosis, blood-brain barrier leakage, and interstitial edema, producing a comparatively uniform ischemic-gliotic signature (McAleese et al., 2017; Hase et al., 2018; Kalaria, 2016; Wardlaw et al., 2019; Joutel & Chabriat, 2017). In AD, lesions reflect a more mixed substrate arising from at least three partially independent processes: coexisting small vessel disease and cerebral amyloid angiopathy (Erten-Lyons et al., 2013; Shirzadi et al., 2023), direct amyloid- and tau-related injury to oligodendrocytes (Lee et al., 2004; Depp et al., 2023; Maitre et al., 2023), and Wallerian-like degeneration secondary to cortical neurodegeneration (McAleese et al., 2017; McAleese et al., 2021). Because these processes need not co-occur in the same proportions across individuals, AD would be expected to yield more heterogeneous lesion signatures than VD. Importantly, the clinical AD/VD distinction is rarely binary; most older adults harbor mixed pathology (Schneider et al., 2007; Toledo et al., 2013; Kapasi et al., 2017), so our analyses are best interpreted as describing the predominant pathological signal in each diagnostic group. These divergent pathways are illustrated in Figure 1.

**Figure 1.**
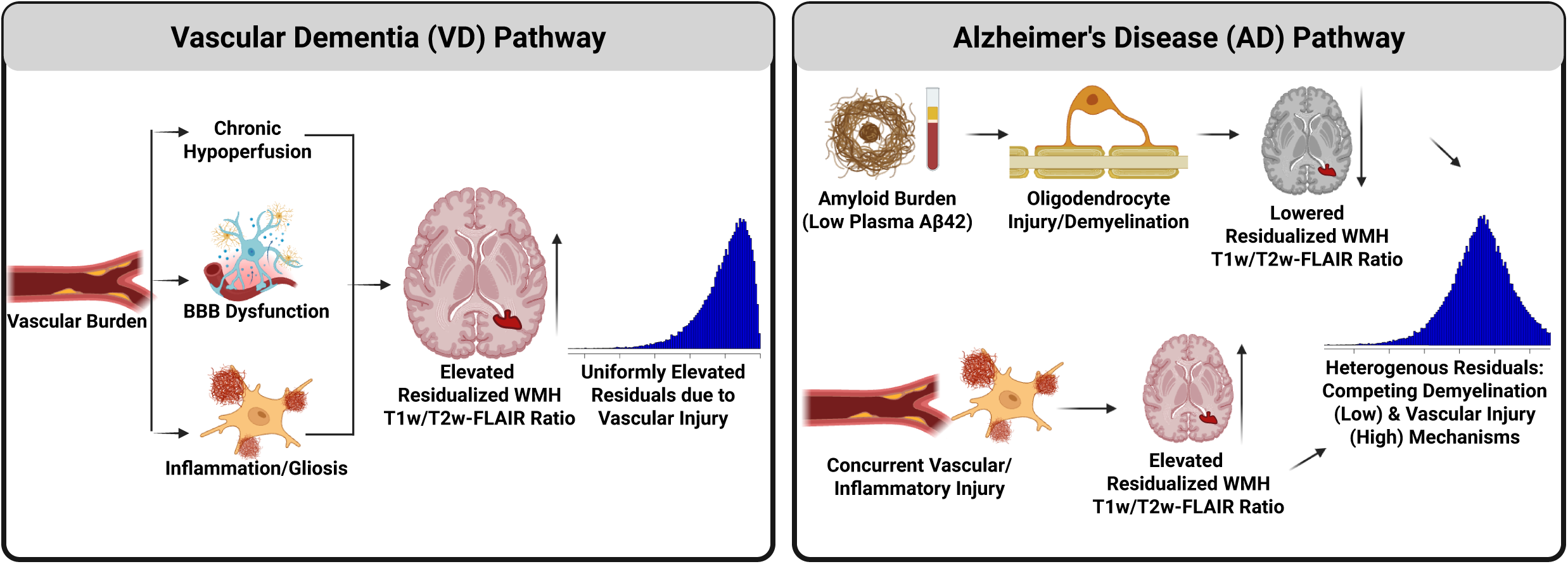
Conceptual model of divergent mechanisms underlying WMH microstructural signatures in vascular dementia and Alzheimer’s disease. The residualized WMH T1w/T2w-FLAIR ratio reflects lesion-specific microstructural deviation after accounting for each individual’s NAWM baseline. Higher residuals indicate WMH ratios exceeding the NAWM-predicted value (interpreted as relative gliotic/cellular response), whereas lower residuals indicate reduced WMH ratios relative to NAWM (interpreted as relative demyelination or tissue rarefaction). Left (VD pathway): Chronic small-vessel disease and hypoperfusion promote blood–brain barrier (BBB) dysfunction and gliotic tissue responses, producing consistently elevated residualized ratios consistent with vascular injury. Right (AD pathway): Greater amyloid burden (lower plasma Aβ42) contributes to oligodendrocyte injury and demyelination, lowering WMH T1w/T2w-FLAIR residuals, while concurrent vascular mechanisms can elevate residuals. The coexistence of amyloid-linked demyelination (lowering) and vascular injury (elevating) yields heterogeneous residualized WMH T1w/T2w-FLAIR profiles in AD. Abbreviations: AD, Alzheimer’s disease; VD, vascular dementia; WMH, white matter hyperintensities; NAWM, normal-appearing white matter; BBB, blood–brain barrier.

A central question is whether microstructural differences in WMH across clinical diagnostic groups reflect AD-related molecular burden rather than purely vascular mechanisms. Cerebral small vessel disease and AD pathology frequently co-occur, motivating investigation of whether WMH-related microstructural differences are mechanistically linked to AD pathology (Sargurupremraj et al., 2020). We therefore aimed to determine whether lesion-specific T1w/T2w-FLAIR ratio differs across the cognitive continuum and whether any such differences are mediated by amyloid burden or by vascular comorbidity. To do so, we leveraged the Biobank Innovations for Chronic Cerebrovascular Disease with Alzheimer’s Disease Study (BICWALZS), which integrates multi-site MRI with AD markers (amyloid PET, plasma p-tau217, NfL, GFAP, Aβ40, Aβ42, and APOE genotyping) alongside clinical phenotyping spanning subjective cognitive impairment (SCI), mild cognitive impairment (MCI), AD, and VD.

Because WMH are hypointense on T1w and hyperintense on T2w-FLAIR, the absolute T1w/T2w-FLAIR ratio within WMH is lower than in normal-appearing white matter (NAWM), and its magnitude is not directly comparable across individuals or scanners. We therefore residualized the WMH T1w/T2w-FLAIR ratio against each participant’s NAWM ratio, yielding a within-scan index of lesion-specific microstructural deviation, and additionally applied ComBat harmonization (see Methods). Two processes move this residual in opposite directions: reactive gliosis and increased cellularity raise the lesion T1w signal disproportionately, producing residuals above the NAWM-predicted value, whereas demyelination and tissue rarefaction lower the ratio, producing residuals below it. We therefore hypothesized that VD would show elevated residuals relative to SCI, reflecting a predominantly ischemic-gliotic phenotype, whereas AD would show more variable residuals, with amyloid-driven demyelination lowering the ratio in some individuals and coexisting vascular injury elevating it in others.

## 2. Methods

### 2.1. Study Design and Participants

Data was obtained from the Biobank Innovations for Chronic Cerebrovascular Disease with Alzheimer’s Disease Study (BICWALZS), a multicenter study initiated in October 2016 by the Korea Disease Control and Prevention Agency to facilitate research on chronic cerebrovascular disease, AD, and mixed pathology. Seven university-affiliated hospitals and community geriatric centers in South Korea participated in data collection, with participants voluntarily recruited from neurology or psychiatry memory outpatient clinics presenting with cognitive concerns. BICWALZS is registered with the Korean National Clinical Trial Registry (KCT0003391), and the study protocol was approved by the Institutional Review Board of Ajou University Hospital (AJOUIRB-SUR-2021-038) and all participating institutions. Participants were recruited voluntarily from those who visited neurology or psychiatry memory out-patient clinics. The original goal was to facilitate, regulate, and ensure optimal use of human biological specimens for research from real-world data.

For this analysis, we included participants who met the following criteria: (1) completion of baseline clinical assessment and neuropsychological testing; (2) 3T brain MRI performed (we included scans within 182 days of clinical evaluation); (3) complete T1-weighted and FLAIR sequences suitable for T1w/T2w-FLAIR ratio calculation; and (4) available plasma biomarker data. We excluded participants with: (1) non-baseline visits; (2) missing T1w/T2w-FLAIR ratio data in WMH regions; or (3) plasma p-tau217 values exceeding 3 pg/mL, which represented statistical outliers based on visual inspection (n=1 excluded; cutoff of 3 pg/mL is 15.17 SD from the mean).

### 2.2. Clinical Diagnosis and Cognitive Assessment

Participants were classified into diagnostic groups using standardized clinical criteria and Clinical Dementia Rating (CDR) staging. SCI was defined as self- and/or informant-reported cognitive complaints in the absence of objective cognitive impairment (no domain score below −1.5 SD) and with preserved daily functioning, corresponding to a global CDR score of 0 (Karim et al., 2022). Mild cognitive impairment (MCI) was diagnosed using the expanded Mayo Clinic criteria, with required evidence of cognitive decline and a global CDR score of 0.5 (Karim et al., 2022; Winblad et al., 2004). Participants were further classified as amnestic or non-amnestic MCI based on memory domain performance (Roh et al., 2022). AD was diagnosed according to the National Institute on Aging-Alzheimer’s Association core clinical criteria for probable AD dementia (Karim et al., 2022; McKhann et al., 2011). VD, specifically subcortical VD, was diagnosed using the major vascular neurocognitive disorder criteria per the Diagnostic and Statistical Manual of Mental Disorders, Fifth Edition (Regier et al., 2013), in conjunction with above-moderate WMH burden on MRI (Karim et al., 2022).

### 2.3. MRI Acquisition

MRI was acquired on 3 Tesla systems from three manufacturers: GE Healthcare (Discovery MR750w), Siemens Healthineers (TrioTim), and Philips Healthcare (Achieva) (Roh et al., 2022, Karim et al., 2022). While scanner models and acquisition parameters varied slightly across the seven clinical sites, all sites adhered to a standardized imaging protocol designed to ensure data compatibility.

Three-dimensional T1-weighted images were acquired using magnetization-prepared rapid gradient echo (MPRAGE) sequences with the following parameters: acquisition matrix 256×256 to 512×512, voxel size 0.39–1.0 mm isotropic, repetition time (TR) 7.1–14.0 ms, echo time (TE) 2.7–6.0 ms, flip angle 8–12°, and slice thickness 1.0–1.3 mm. Fluid-attenuated inversion recovery (FLAIR) images were acquired with acquisition matrix 224×256 to 512×560, voxel size 0.39–0.94 mm in-plane, TR 8.8–12.0 s, TE 80–146 ms, and slice thickness 2.0–7.0 mm (Roh et al., 2022; Karim et al., 2022). Detailed sequence parameters for each site are provided elsewhere (Roh et al., 2022; Karim et al., 2022).

All MR scans underwent visual quality assurance before processing. Scans were excluded for substantial motion artifact, incomplete brain coverage, or major structural abnormalities (e.g., tumor, large stroke, prior surgical resection). To further check that bias-field correction and intensity normalization produced a consistent signal across the cohort, we computed voxel-wise minimum and maximum intensity maps across scans and visually inspected them for outliers.

### 2.4. White Matter Hyperintensity Segmentation

White matter hyperintensities were identified and segmented using the Lesion Segmentation Toolbox with artificial intelligence (LST-AI) algorithm for detecting white matter lesions on FLAIR sequences (Wiltgen et al., 2024). This employs an ensemble of three 3D U-Net models that process skull-stripped and intensity-normalized images. A final lesion map is generated by thresholding three lesion probability maps at 0.5. All automated segmentations underwent visual quality control by trained analysts who were blinded to participants’ clinical diagnoses.

We conducted FreeSurfer v7.1.1 on T1-weighted MPRAGE that were bias corrected (Fischl, 2012). This pipeline conducts motion correction, removal of non-brain tissue, automated coregistration to standardized spaces, intensity normalization, tessellation of gray matter/white matter boundaries, and optimization of borders of gray/white matter boundaries. Normal-appearing white matter (NAWM) was defined as white matter tissue that was classified as white matter by FreeSurfer and not identified by LST-AI as WMH.

### 2.5. T1w/T2w-FLAIR Ratio Calculation and Residualization

Because no conventional T2-weighted sequence was available in BICWALZS, we used the FLAIR sequence as the T2-weighted image throughout the ratio calculation; we therefore refer to this measure as the T1w/T2w-FLAIR ratio. Unlike a traditional T2-weighted sequence, the FLAIR is largely a T2-weighted contrast. We estimated the T1w/T2w-FLAIR ratio using a custom singularity container (https://github.com/tetra-tools/T1_T2_Ratio) adapted from a previously established method (https://github.com/treanus/kul_nis). These tools implement the approach described by Ganzetti et al. (2014) and Cappelle et al. (2022). First, T2-weighted images were rigidly co-registered to the corresponding T1-weighted images using Advanced Normalization Tools (ANTs, version 2.3.1) (Avants et al., 2009). Both T1-weighted and co-registered T2-weighted images underwent bias field correction to minimize intensity inhomogeneities. We then conducted normalization to MNI space using antsRegistrationSyN (affine + rigid + nonlinear; Ganzetti et al., 2014) and conducted brain extraction using HD-BET (Isensee et al., 2019). We inverse normalized an extra-cerebral mask to native space to extract an intensity histogram of these tissues for the T1w images and MNI space T1w template images. We conducted nonlinear histogram matching using mrhistmatch (Tournier et al., 2019) that was applied to the T1w image relative to an existing MNI space T1w image. This entire process was similarly done for the T2w FLAIR images. This aligns the full T1w and FLAIR intensity distributions to a common reference, controlling the overall distribution shape rather than only the mean. We then calculate voxel-wise T1w/T2w-FLAIR ratio using the MPRAGE divided by the FLAIR. This procedure is detailed in Capelle et al. (Cappelle et al., 2022) and has been shown to effectively estimate T1w/T2w-FLAIR ratio.

We then extracted the mean T1w/T2w-FLAIR ratio in the WMH and NAWM in native space. To isolate lesion-specific microstructural deviations from each participant’s own white matter baseline, we regressed the WMH T1w/T2w-FLAIR ratio against the NAWM T1w/T2w-FLAIR ratio (model: WMH T1w/T2w-FLAIR ∼ NAWM T1w/T2w-FLAIR + residual) and used standardized residuals (hereafter ‘T1w/T2w-FLAIR ratio residuals’) as the primary outcome. Conceptually, this is equivalent to expressing each participant’s lesion signal relative to their own NAWM signal – the same approach as normalizing WMH T1w/T2w-FLAIR by NAWM T1w/T2w-FLAIR – but it has the additional advantage of preserving the linear scale of the original measure for downstream regression. This within-scan, within-individual referencing further reduces residual interindividual, site, and scanner-related variability that persists despite intensity normalization and isolates WMH-specific microstructural shifts.

### 2.6. Amyloid PET Acquisition and Quantification

Participants underwent brain amyloid imaging using 18F-flutemetamol positron emission tomography (PET) on Discovery STE or Discovery 690 PET/CT scanners (GE Healthcare). 18F-flutemetamol (mean dose 185 MBq) was administered as an intravenous bolus (Roh et al., 2022). Following a 90-minute uptake period, a 20-minute static PET acquisition was performed, consisting of four 5-minute frames (Roh et al., 2022). PET images were co-registered to T1-weighted MRI using rigid-body transformation, normalized to standard template space, averaged across frames, and quantified as standardized uptake value ratios (SUVRs) using the pons as reference region (Roh et al., 2022). All preprocessing was done using SPM12.

Global cortical 18F-flutemetamol retention was quantified as the volume-weighted average SUVR across 28 bilateral cortical regions of interest derived from the automated anatomical labeling (AAL) atlas, including the frontal, posterior cingulate, lateral temporal, parietal, and occipital lobes. Cortical SUVR values were converted to Centiloid units using the equation: Centiloid = 100 × (SUVR −0.634) / (1.548 - 0.634), where 0.634 and 1.548 represent the mean SUVR values for amyloid-negative and amyloid-positive reference populations, respectively (Klunk et al., 2015; Roh et al., 2022).

Participants were classified as amyloid-positive if their Centiloid value was greater than or equal to 30, corresponding to an SUVR threshold of approximately 0.65 (Roh et al., 2022; Karim et al., 2022). This threshold has been validated in previous studies of older Korean adults and aligns with established cutoffs for 18F-flutemetamol (Roh et al., 2022).

### 2.7. Plasma Biomarker Quantification

Blood samples were taken after an overnight fast in the morning by venipuncture and collected in serum separating tubes and dipotassium ethylenediamineetetraacetic acid. Blood samples were stabilized and centrifuged at 3000rpm for 10min at room temperature to obtain plasma and serum supernatants. Plasma and serum supernatants were further centrifuged under the same conditions and stored in a –80C deep freezer. Plasma biomarkers were measured using Single Molecule Array (Simoa) platform (Quanterix®) and performed on Simoa HD-X Analyzer at the University of Pittsburgh. The following biomarkers were quantified: amyloid-β 42 (Aβ42), amyloid-β 40 (Aβ40), glial fibrillary acidic protein (GFAP), and neurofilament light chain (NfL) (Neurology 4-Plex E assay, #103670) and, p-tau217 (ALZpath; Ashton et al., 2024). All assays were performed according to manufacturer protocols by laboratory personnel blinded to participants’ clinical diagnoses.

### 2.8. *APOE* Genotyping

Informed consent was obtained from all participants for collection and genotyping of blood genomic DNA. Genomic DNA was isolated from blood samples, and single nucleotide polymorphism (SNP) genotyping was performed by DNA Link, Inc. (Seoul, Korea) using the Affymetrix Axiom KORV1.0-96 Array (Thermo Fisher Scientific) according to manufacturer protocol (Roh et al., 2022). *APOE* genotypes were derived from rs429358 and rs7412, which define the *APOE* ε2, ε3, and ε4 alleles. Quality control procedures included assessment of call rates (threshold ≥97%), Hardy-Weinberg equilibrium testing, and verification of gender concordance (Roh et al., 2022).

Participants were classified into three *APOE* groups based on their genotype: (1) E2 carriers – individuals carrying at least one ε2 allele without ε4 (E2/E2, E2/E3); (2) E3 homozygotes – homozygous for the ε3 allele (E3/E3); and (3) E4 carriers – individuals carrying at least one ε4 allele (E2/E4, E3/E4, E4/E4).

### 2.9. Statistical Analysis

All statistical analyses were conducted using R v.4.4.1. To address data missingness in plasma biomarkers and covariates, we employed Multiple Imputation by Chained Equations (MICE) using the Random Forest method. Five imputed datasets were generated, and parameter estimates from all subsequent regression and mediation models were pooled according to Rubin’s rules. To verify the stability of our findings, we performed parallel sensitivity analyses using non-imputed, complete-case data; any notable divergences between the primary imputed results and complete-case analyses are reported, though results were convergent.

We assessed the biological and clinical determinants of T1w/T2w-FLAIR ratio residuals using a series of linear regression models. To isolate the effects of pathology from physiological aging and demographic confounders, all models included age, sex, years of education, study site, and the interval between MRI and clinical assessment as covariates. We modeled T1w/T2w-FLAIR ratio residuals as the dependent variable against predictors, including clinical diagnostic group (SCI as the reference), *APOE* ε4 carrier status (with E2/E2, E2/E3 as the reference), amyloid-PET positivity (negative as the reference), and specific plasma biomarkers (p-tau217, Aβ42, Aβ40, GFAP, and NfL). We additionally calculated plasma Aβ42/Aβ40 and p-tau217/Aβ42 ratios. Before analysis, these derived ratios were normalized using the bestNormalize package in R, which applies the optimal transformation to achieve a Gaussian distribution – in both cases it used rank transformation.

To investigate whether specific biological mechanisms link diagnosis to white matter microstructural deviations, we conducted causal mediation analyses using Structural Equation Modeling (SEM) via the lavaan package. We constructed models to test whether the effect of an AD diagnosis on the T1w/T2w-FLAIR ratio was mediated through amyloid pathology (indexed by plasma Aβ40 and Aβ42). These were constrained to these two since we found associations with those two markers (see Section 3.3). These models decomposed the total effect of diagnosis into: (1) path *a*, the effect of diagnosis on the biomarker; (2) path *b*, the effect of the biomarker on the T1w/T2w-FLAIR ratio; and (3) path *c’*, the direct effect of diagnosis on the T1w/T2w-FLAIR ratio independent of the biomarker. The indirect effect was calculated as the product of coefficients (axb), and the proportion mediated was derived by dividing the indirect effect by the total effect (axb+c’). We additionally tested this mediation for VD.

To examine the potential contribution of vascular comorbidity to white matter microstructure, we constructed a composite vascular risk score from eight binary clinical diagnoses: hypertension, dyslipidemia, diabetes, cerebrovascular disease, peripheral vascular disease, myocardial infarction, congestive heart failure, and angina. Each condition was recorded as present or absent and summed to yield a total score. Participants were categorized into four groups (0, 1, 2, or ≥3 risk factors). As a sensitivity analysis, this vascular risk category was added as an additional covariate to the primary linear regression model examining plasma Aβ42 as a predictor of T1w/T2w-FLAIR ratio residuals, with age, sex, years of education, study site, diagnostic group, and MRI-to-clinical interval retained as covariates. This model was estimated across all imputed datasets with pooled inference. To further investigate vascular mechanisms within the VD group, we additionally tested a mediation model in which VD diagnosis served as the independent variable, high vascular burden (≥3 risk factors, coded as binary) as the mediator, and T1w/T2w-FLAIR ratio residuals as the outcome, with plasma Aβ42 and the same demographic and site covariates included as adjustments throughout. This was done as this group showed differences in T1w/T2w-FLAIR ratio (see Section 3.3).

Across this study, we conducted 12 independent linear regression models examining predictors of the residualized T1w/T2w-FLAIR ratio. To account for multiple comparisons across these models, we applied a Bonferroni correction to the primary multivariable model (Table 2). Under this, a p-value is considered significant after correction if it is below 0.05/12 = 0.0042. Corrected p-values for the primary model are reported in Section 3.3.

Moreover, since participating sites differed in scanner manufacturer and acquisition parameters, we performed a sensitivity analysis using ComBat harmonization (Fortin et al., 2017; Johnson et al., 2007) on the NAWM and WMH T1w/T2w-FLAIR ratios before residualization. ComBat models site as a batch variable while preserving biological covariates of interest (age, sex, education, days between scan and visit, and clinical diagnosis). We refit the primary multivariable model (Table 2) on the harmonized data and compared the resulting coefficients and significance with the unharmonized analysis. The harmonized model results are presented in Supplemental Table 13.

## 3. Results

### 3.1. Participant Characteristics

The sample analyzed included 576 participants (from an initial cohort of 668). Inclusion criteria required a baseline clinical diagnosis, complete T1w/T2w-FLAIR ratio, a maximum interval of 182 days between imaging and clinical assessment, and any outliers in biological variables. The cohort contained individuals with SCI (N=71), MCI non-amnestic (N=74), MCI amnestic (N=196), AD (N=125), VD (N=88), and other disorders/dementias (N=22). Participants were recruited across seven clinical sites in South Korea (Table 1).

**Table 1.** Demographic and Clinical Characteristics by Diagnostic Group (N = 576)

| Characteristic | Overall<br>(N = 576) | SCI<br>(N = 71) | MCI non-<br>amnesic<br>(N = 74) | MCI<br>amnesic<br>(N = 196) | AD<br>(N = 125) | VD<br>(N = 88) | Other<br>(N = 22) |
| --- | --- | --- | --- | --- | --- | --- | --- |
| Age (years), mean (SD) | 72.4 (7.6) | 71.3 (6.8) | 72.0 (7.3) | 71.8 (7.5) | 73.6 (8.5) | 73.6 (7.5) | 71.4 (8.1) |
| Sex, n (%) |  |  |  |  |  |  |  |
| Female | 385 (66.8) | 53 (74.6) | 57 (77.0) | 130 (66.3) | 76 (60.8) | 56 (63.6) | 13 (59.1) |
| Male | 191 (33.2) | 18 (25.4) | 17 (23.0) | 66 (33.7) | 49 (39.2) | 32 (36.4) | 9 (40.9) |
| Education (years), mean (SD) | 8.2 (4.7) | 7.1 (3.9) | 7.2 (4.7) | 9.1 (4.9) | 8.9 (4.6) | 7.2 (4.9) | 6.2 (3.7) |
| Days between scan and visit,<br>mean (SD) | 17.9 (31.1) | 36.0 (46.8) | 7.2 (12.9) | 19.9 (32.4) | 16.3 (29.9) | 12.8 (20.1) | 7.0 (11.8) |
| Site, n (%) |  |  |  |  |  |  |  |
| Ajou | 170 (29.5) | 17 (23.9) | 19 (25.7) | 59 (30.1) | 44 (35.2) | 27 (30.7) | 4 (18.2) |
| Chonnam | 18 (3.1) | 0 (0.0) | 1 (1.4) | 2 (1.0) | 8 (6.4) | 4 (4.5) | 3 (13.6) |
| Gwangju | 28 (4.9) | 19 (26.8) | 2 (2.7) | 4 (2.0) | 1 (0.8) | 2 (2.3) | 0 (0.0) |
| Inha | 111 (19.3) | 18 (25.4) | 11 (14.9) | 45 (23.0) | 24 (19.2) | 9 (10.2) | 4 (18.2) |
| Pusan | 27 (4.7) | 4 (5.6) | 1 (1.4) | 8 (4.1) | 11 (8.8) | 1 (1.1) | 2 (9.1) |
| Samsung | 87 (15.1) | 2 (2.8) | 2 (2.7) | 45 (23.0) | 26 (20.8) | 12 (13.6) | 0 (0.0) |
| Suwon | 135 (23.4) | 11 (15.5) | 38 (51.4) | 33 (16.8) | 11 (8.8) | 33 (37.5) | 9 (40.9) |
| APOE Genotype, n (%) |  |  |  |  |  |  |  |
| E2 carrier | 66 (11.5) | 9 (12.7) | 10 (13.5) | 24 (12.2) | 4 (3.2) | 14 (15.9) | 5 (22.7) |
| E3 carrier | 334 (58.0) | 53 (74.6) | 45 (60.8) | 113 (57.7) | 55 (44.0) | 57 (64.8) | 11 (50.0) |
| E4 carrier | 176 (30.6) | 9 (12.7) | 19 (25.7) | 59 (30.1) | 66 (52.8) | 17 (19.3) | 6 (27.3) |
| Amyloid PET Status, n (%) |  |  |  |  |  |  |  |
| Negative | 308 (53.5) | 52 (73.2) | 55 (74.3) | 94 (48.0) | 20 (16.0) | 69 (78.4) | 18 (81.8) |
| Positive | 185 (32.1) | 7 (9.9) | 12 (16.2) | 60 (30.6) | 92 (73.6) | 12 (13.6) | 2 (9.1) |
| Missing | 83 (14.4) | 12 (16.9) | 7 (9.5) | 42 (21.4) | 13 (10.4) | 7 (8.0) | 2 (9.1) |
| Plasma p-tau217, mean (SD) | 0.1 (0.1) | 0.1 (0.1) | 0.1 (0.1) | 0.1 (0.1) | 0.3 (0.2) | 0.1 (0.1) | 0.1 (0.1) |
| Plasma Aβ42, mean (SD) | 4.0 (2.0) | 3.9 (2.4) | 3.8 (1.7) | 4.0 (1.7) | 3.5 (2.0) | 4.6 (2.5) | 4.1 (1.9) |
| Plasma Aβ40, mean (SD) | 64.4 (34.9) | 62.8 (37.6) | 61.4 (26.5) | 66.8 (27.0) | 58.9 (35.2) | 72.5 (50.7) | 56.4 (32.3) |
| Plasma GFAP, mean (SD) | 186.0 (129.5) | 131.4<br>(67.8) | 145.2 (72.1) | 184.8 (155.5) | 250.0<br>(107.5) | 178.6<br>(132.0) | 166.7<br>(136.8) |
| Plasma NFL, mean (SD) | 37.1 (30.6) | 29.2 (31.1) | 28.6 (14.5) | 34.2 (32.2) | 46.4 (29.8) | 41.1 (30.6) | 46.5 (40.9) |

**Abbreviations:** AD, Alzheimer's disease; APOE, apolipoprotein E; Aβ , amyloid-beta; GFAP, glial fibrillary acidic protein; MCI, mild cognitive impairment; NFL, neurofilament light; PET, positron emission tomography; SCI, subjective cognitive impairment; SD, standard deviation; VD, vascular dementia.
**Note:** Percentages for amyloid PET status calculated from available data (N = 495 with PET data). Plasma biomarker sample sizes vary by marker due to missing data.

**Table 2.** Association of Residualized T1w/T2w-FLAIR Ratio with Plasma A. β**42**

| Term | Standardized $\beta$ | Standard Error | P-value | Standardized $\beta$ (Imputed) | P-value (Imputed) |
| --- | --- | --- | --- | --- | --- |
| **Intercept** | **-0.556** | **0.076** | **$\leq 0.001$** | **-0.62** | **$\leq 0.001$** |
| **Age** | **0.175** | **0.001** | **$\leq 0.001$** | **0.15** | **$\leq 0.001$** |
| Sex (Male) | -0.011 | 0.016 | 0.907 | 0.057 | 0.504 |
| Education (years) | 0.027 | 0.002 | 0.577 | 0.006 | 0.883 |
| Days Between Scan and Visit | -0.111 | 0 | 0.061 | -0.078 | 0.189 |
| Site: Chonnam | 0.081 | 0.042 | 0.75 | 0.113 | 0.644 |
| Site: Gwangju | 0.69 | 0.046 | 0.013 | 0.565 | 0.037 |
| **Site: Inha** | **0.281** | **0.022** | **0.036** | **0.318** | **0.004** |
| Site: Pusan | 0 | 0.034 | 1 | 0.07 | 0.732 |
| **Site: Samsung** | **0.738** | **0.024** | **$\leq 0.001$** | **0.841** | **$\leq 0.001$** |
| Site: Suwon | 0.016 | 0.02 | 0.896 | 0.136 | 0.222 |
| Group: MCI non-amnestic | 0.047 | 0.03 | 0.792 | 0.057 | 0.729 |
| Group: MCI amnestic | 0.214 | 0.026 | 0.167 | 0.204 | 0.133 |
| **Group: AD** | **0.431** | **0.028** | **0.012** | **0.374** | **0.011** |
| **Group: VD** | **0.304** | **0.029** | **0.086** | **0.311** | **0.04** |
| Group: Other | 0.181 | 0.043 | 0.485 | 0.182 | 0.42 |
| **Plasma Aβ42** | **0.1** | **0.004** | **0.029** | **0.093** | **0.054** |
| Vascular Risk: 1 | 0.075 | 0.019 | 0.52 | 0.065 | 0.558 |
| Vascular Risk: 2 | 0.15 | 0.021 | 0.226 | 0.17 | 0.122 |
| **Vascular Risk: 3+** | **0.445** | **0.023** | **0.001** | **0.437** | **0.001** |

**Abbreviations:** AD, Alzheimer's disease; Aβ 42, amyloid-beta 42; MCI, mild cognitive impairment; SCI, subjective cognitive impairment; VD, vascular dementia; CI, confidence interval.
**Notes:** Model adjusted for age, sex, education, site, cognitive diagnosis, days between scan and visit, plasma Aβ 42, and vascular risk category. The dependent variable is the residualized global T1w/T2w-FLAIR ratio (global ratio adjusted for normal-appearing white matter). Plasma Aβ 42 was modeled as a continuous variable; vascular risk was modeled categorically (0 as reference; levels 1, 2, and $\geq 3$ shown). Reference categories: Group = SCI; Sex = Female; Site = Ajou. Linear regression with residualized T1w/T2w-FLAIR ratio as the dependent variable. CI = 95% confidence interval. All estimates represent standardized regression coefficients ( $\beta$ ). P-values $\leq 0.001$ are denoted as $\leq 0.001$ .

### 3.2. T1w/T2w-FLAIR Ratio Residualization

The global T1w/T2w-FLAIR ratio was strongly correlated with the NAWM T1w/T2w-FLAIR ratio (B=1.03, SE=0.014, t=76.13, p<2×10 ¹), with 91% of the variance (R^2^) in the global signal being attributable to white matter contributions. To isolate lesion-specific microstructural pathology from this global baseline, we calculated the T1w/T2w-FLAIR ratio residuals. This ratio residual represents the deviation of tissue integrity within white matter hyperintensities (WMH) relative to the individual’s normal-appearing white matter (NAWM), serving as the primary outcome for all subsequent analyses. By normalizing WMH signal to each participant’s NAWM baseline, this residualized measure minimizes external effects and focuses inference on lesion-specific microstructural deviation. Higher residuals indicate WMH T1w/T2w-FLAIR ratios that exceed each participant’s NAWM-predicted value, interpreted as relative tissue preservation, gliotic/cellular response, or reduced edema. Whereas lower residuals indicate WMH ratios falling below the NAWM-predicted value, interpreted as relative demyelination or tissue rarefaction. Since the residual is computed relative to each participant’s own NAWM signal, it represents the deviation of the lesion intensity from that individual’s internal reference and is conceptually equivalent to expressing the WMH T1w/T2w-FLAIR ratio as a fraction of NAWM T1w/T2w-FLAIR, while preserving the linear scale of the original measure.

### 3.3. Differences in T1w/T2w-FLAIR Ratio

Models using pooled imputed data and not imputed data resulted in similar regression coefficients – we present both in all tables. We found that greater age was associated with greater T1w/T2w-FLAIR ratio (Supplemental Table 1, Figure 2A) but not sex (Supplemental Figure 1A). We found that both the VD and AD groups showed heightened T1w/T2w-FLAIR ratio relative to the SCI group (Supplemental Table 2, Figure 2B) but found no differences between the AD and VD groups. We found that lower T1w/T2w-FLAIR ratios were associated with lower plasma Aβ42 (Supplemental Table 3, Figure 2C) and Aβ40 (Supplemental Table 4) (i.e., greater brain amyloid burden).

**Figure 2.**
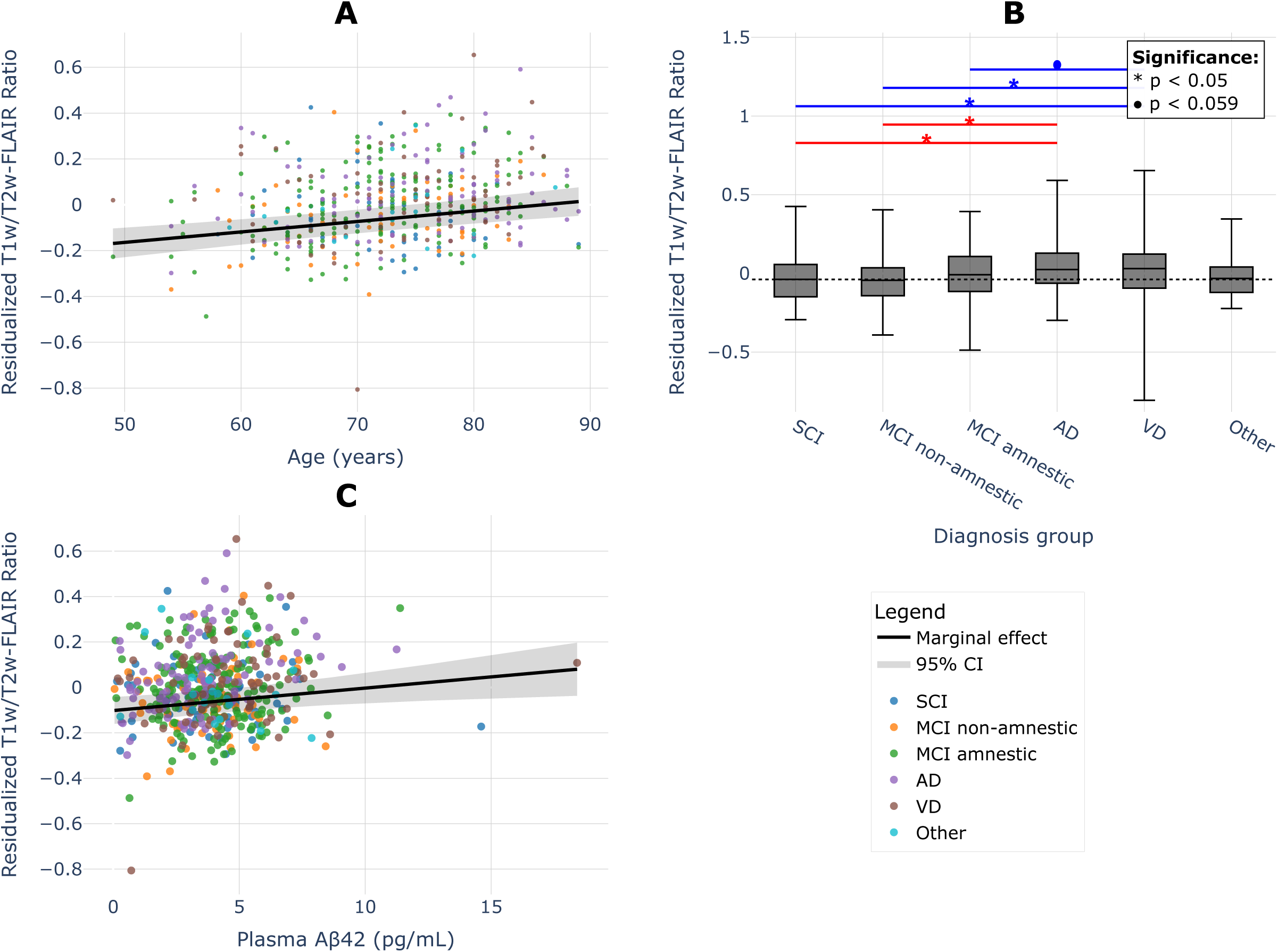
Association of residualized T1w/T2w-FLAIR ratio with age, cognitive diagnosis, and plasma. **A**β**42. (A)** Scatter plot showing the relationship between age and residualized T1w/T2w-FLAIR ratio across all cognitive diagnostic groups. The black line represents the marginal effect of age (β=0.004, p<0.001) with 95% confidence interval (gray shaded area), adjusted for sex, education, site, cognitive diagnosis, and days between scan and visit. **(B)** Boxplot showing residualized T1w/T2w-FLAIR ratio (adjusted for covariates) across cognitive diagnostic groups. The dotted horizontal line represents the median residualized T1w/T2w-FLAIR ratio for the SCI group. Asterisks denote diagnostic groups with significantly higher residualized T1w/T2w-FLAIR ratios compared with the SCI group (p < 0.05). **(C)** Scatter plot demonstrating the association between plasma Aβ42 concentration and residualized T1w/T2w-FLAIR ratio across diagnostic groups. The black line shows the marginal effect of Aβ42 (β=0.0099, p=0.009) with 95% confidence interval (gray shaded area), adjusted for age, sex, education, site, cognitive diagnosis, and days between scan and visit. All panels display individual data points colored by cognitive diagnostic group: SCI (subjective cognitive impairment), MCI Non-Amnestic (mild cognitive impairment non-amnestic), MCI Amnestic (mild cognitive impairment amnestic), AD (Alzheimer’s disease), VD (vascular dementia), and other diagnoses. The data points visible at the upper end of the plasma Aβ42 distribution in panel C are not statistical outliers - they were retained after pre-specified exclusion criteria (which removed only p-tau217 values exceeding 3 pg/mL, defined a priori) and represent biologically plausible plasma Aβ42 values that fall within the published reference range for the Quanterix Simoa N4PE assay in older adults. Sensitivity inspection confirmed that excluding these participants did not alter the direction or significance of the Aβ42–T1w/T2w-FLAIR association.

We found no association between T1w/T2w-FLAIR ratio and *APOE* genotype (Supplemental Table 5, Supplemental Figure 1B), amyloid PET status (Supplemental Table 6), p-tau217 (Supplemental Table 7), NfL (Supplemental Table 8), or GFAP (Supplemental Table 9). We additionally investigated the association of composite plasma biomarkers with the T1w/T2w-FLAIR ratio. We found no significant association with the Aβ42/40 ratio (Supplemental Table 10) or the p-tau217/Aβ42 ratio (Supplemental Table 11).

As a sensitivity analysis, we added vascular risk categories to the primary plasma Aβ42 model (Table 2). Greater vascular comorbidity burden was independently associated with higher T1w/T2w-FLAIR ratio residuals, with ≥3 risk-factor category showing a robust positive association in both complete-case and imputed analyses. The associations with AD and VD diagnosis, as well as plasma Aβ42, remained directionally consistent after adjusting for vascular burden (Table 2).

Consequently, our primary multivariable model focused on plasma Aβ42 alongside clinical diagnosis (Table 2). In this model, greater age was associated with a greater T1w/T2w-FLAIR ratio, and both the VD and AD groups exhibited heightened T1w/T2w-FLAIR ratios. Of these, the associations with age and with ≥3 vascular risk factors survived Bonferroni correction across the 12 hierarchical models we conducted (α = 0.0042). The AD and VD group effects and the plasma Aβ42 association did not reach this threshold and should be interpreted as nominally significant.

We conducted ComBat-harmonized sensitivity analysis and this yielded coefficient estimates and significance for the tested variables (age, AD and VD diagnosis, plasma Aβ42, and ≥3 vascular risk factors) that were materially equivalent to the primary unharmonized model (Supplemental Table 13). As expected, the site coefficients were substantially attenuated after harmonization, confirming that the procedure removed the targeted between-site variation. These results indicate that the reported associations are not driven by between-site scanner differences.

Given the significant associations between AD diagnosis, plasma biomarkers, and the T1w/T2w-FLAIR ratio, we subsequently explored mediation models to evaluate whether Aβ40 or Aβ42 mediated the relationship between AD and T1w/T2w-FLAIR ratio. We repeated this for VD. We similarly conducted mediation analysis with ≥3 risk-factor category as a mediator between VD and T1w/T2w-FLAIR ratio as well as AD and T1w/T2w-FLAIR ratio.

### 3.4. Mediation Analysis

Structural equation modeling showed mechanistic divergence in the pathology of white matter injury between Alzheimer’s and vascular phenotypes. Mediation analyses indicated that the effect of AD diagnosis on the T1w/T2w-FLAIR ratio was partially mediated by plasma Aβ42 (Table 3, Figure 3B).

**Figure 3.**
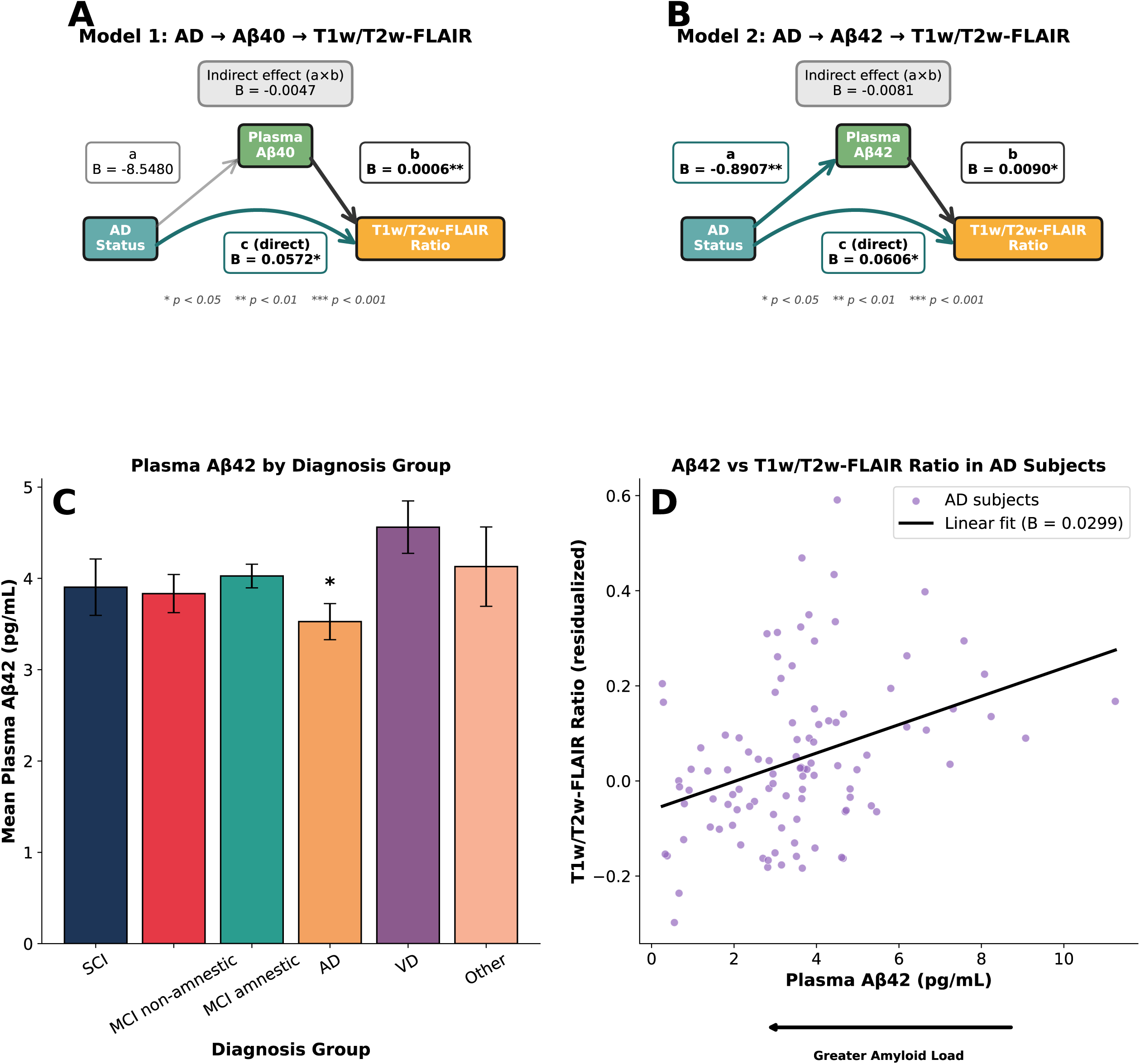
Mediation analysis of Alzheimer’s disease diagnosis on T1w/T2w-FLAIR ratio via plasma amyloid biomarkers. Path diagrams illustrating mediation analyses examining whether the effect of AD diagnosis on residualized T1w/T2w-FLAIR ratio is mediated through plasma amyloid biomarkers. (A) Mediation model with plasma Aβ40 as mediator. Path a represents the effect of AD diagnosis on Aβ40 (B = −8.548, p = 0.113), path b represents the effect of Aβ40 on T1w/T2w-FLAIR ratio controlling for AD diagnosis (B = 0.0006, p < 0.01), and path c represents the direct effect of AD diagnosis on T1w/T2w-FLAIR ratio (B = 0.057, p < 0.05). The indirect effect was not significant (a×b = −0.005, p = 0.162). (B) Mediation model with plasma Aβ42 as mediator. Path a shows a significant effect of AD diagnosis on reduced Aβ42 (B = −0.891, p < 0.01), path b shows the effect of Aβ42 on T1w/T2w-FLAIR ratio (B = 0.009, p < 0.05), and path c represents the direct effect (B = 0.061, p < 0.05). The indirect effect showed a partial mediation (a×b = −0.008, p = 0.068). (C) Bar plot showing mean plasma Aβ42 concentrations (pg/mL) across diagnostic groups with error bars representing standard error. (D) Scatter plot of T1w/T2w-FLAIR ratio residuals versus plasma Aβ42 for AD participants, with linear regression fit. All mediation models adjusted for age, sex, education, site, diagnosis group, and days between scan and visit. *p < 0.05, **p < 0.01, ***p < 0.001.

**Table 3.** Mediation Analysis Results.

| Model | Path | Effect | Coefficient (B) | Standard Error | P-value |
| --- | --- | --- | --- | --- | --- |
| Model 1: AD Status → Aβ40 → T1w/T2w-FLAIR Ratio | a (AD → Aβ40) | Direct | -8.54805 | 5.386647 | 0.112852 |
|  | b (Aβ40 → T1w/T2w-FLAIR) | Direct | 0.000555 | 0.000192 | 0.00392 |
|  | c (AD → T1w/T2w-FLAIR) | Direct | 0.057197 | 0.024622 | 0.02038 |
|  | ab | Indirect | -0.00471 | 0.003365 | 0.161507 |
| Model 2: AD Status → Aβ42 → T1w/T2w-FLAIR Ratio | a (AD → Aβ42) | Direct | -0.89066 | 0.288868 | 0.002104 |
|  | b (Aβ42 → T1w/T2w-FLAIR) | Direct | 0.00903 | 0.00383 | 0.018576 |
|  | c (AD → T1w/T2w-FLAIR) | Direct | 0.060567 | 0.024727 | 0.014478 |
|  | ab | Indirect | **-0.00808** | **0.004428** | **0.068185** |

Abbreviations: AD, Alzheimer's disease; SE, standard error; β, Beta coefficient.
Note: Model 1 evaluates whether the association between AD status and residualized T1w/T2w-FLAIR ratio is mediated through plasma Aβ40. Model 2 evaluates whether the association between AD status and residualized T1w/T2w-FLAIR ratio is mediated through plasma Aβ42. All mediation models were adjusted for age, sex, years of education, imaging site, cognitive diagnostic group, and days between MRI acquisition and clinical visit. Path a represents the effect of the predictor on the mediator; path b represents the effect of the mediator on residualized T1w/T2w-FLAIR ratio; path c represents the direct effect of the predictor on residualized T1w/T2w-FLAIR ratio after accounting for the mediator; ab represents the indirect (mediated) effect through the biomarker. Bold values indicate $p < 0.05$ .

Plasma Aβ40 (Figure 3A) did not show a significant mediation effect. We present that Aβ42 shows similar expected differences with AD < SCI < VD, where individuals with AD show low Aβ42 indicative of greater amyloid pathology (Figure 3C), and that a lower T1w/T2w-FLAIR ratio is associated with lower Aβ42 (Figure 3D). We repeated the same analyses for VD but found no mediating effect for either Aβ40 or Aβ42 (Supplemental Table 12).

In VD, vascular comorbidity burden partially mediated the relationship between diagnosis and T1w/T2w-FLAIR ratio. VD diagnosis was significantly associated with high vascular burden (≥3 risk factors), which in turn was associated with greater T1w/T2w-FLAIR ratio, consistent with a partial indirect pathway (Supplemental Table 12, Figure 4A). The indirect effect was significant, and a direct effect of VD on T1w/T2w-FLAIR ratio residuals remained after accounting for vascular, indicating partial mediation rather than full mediation (Supplemental Table 12, Figure 4A). The distribution of vascular risk categories across diagnostic groups and the stepwise increase in T1w/T2w-FLAIR ratio residuals with increasing vascular burden are shown in Figure 4B-C. We found that high vascular burden did not mediate the association between AD and T1w/T2w-FLAIR ratio.

**Figure 4.**
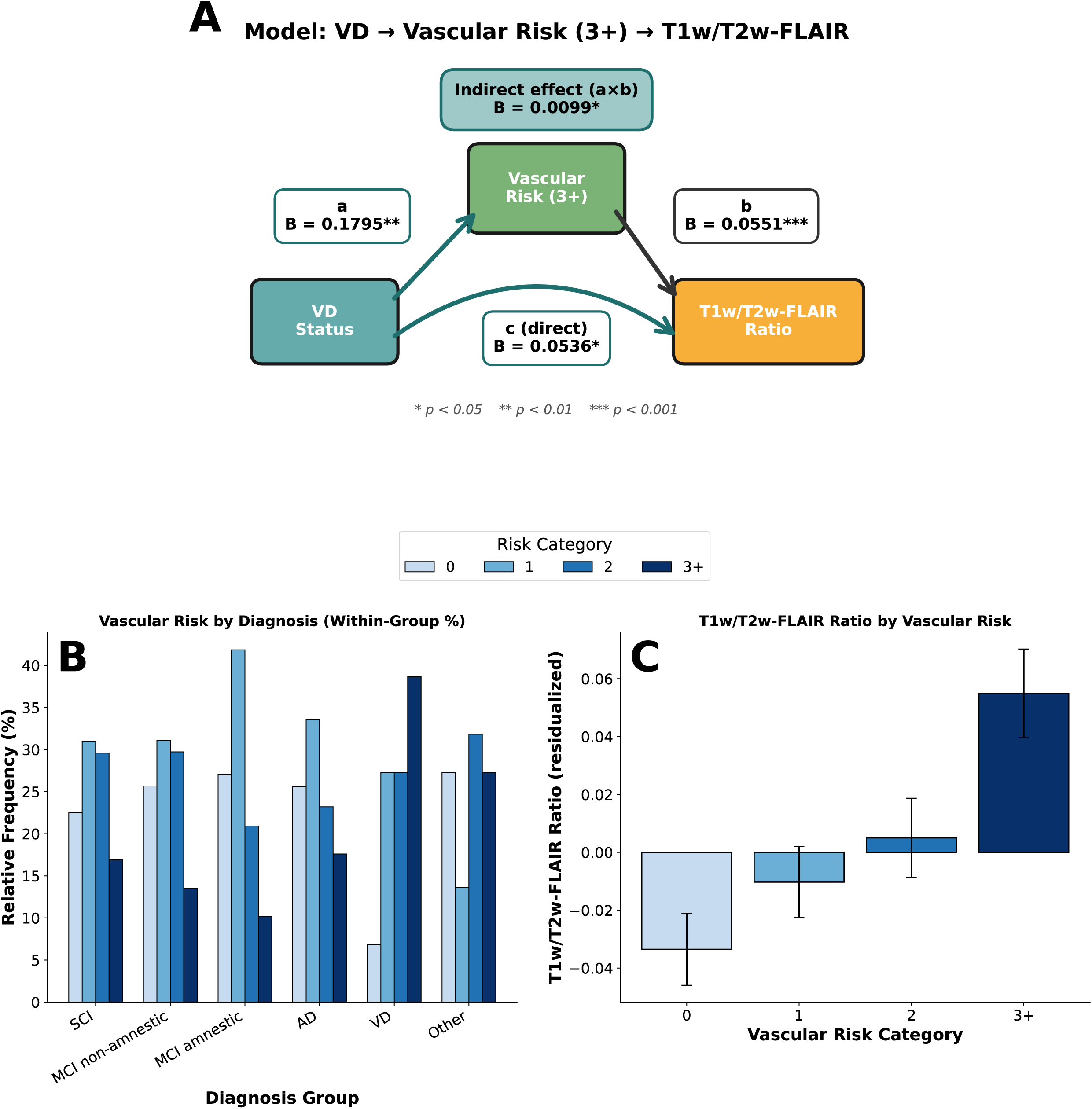
Mediation analysis of vascular dementia diagnosis on T1w/T2w-FLAIR ratio via vascular risk burden. The path diagram and supporting plots show a mediation analysis examining whether high vascular risk burden (≥3 risk factors) mediates the effect of VD diagnosis on residualized T1w/T2w-FLAIR ratio. (A) Mediation model with vascular risk category (3+) as mediator. Path a represents the effect of VD diagnosis on vascular risk burden (B = 0.1795, p < 0.01), path b represents the effect of vascular risk burden on T1w/T2w-FLAIR ratio controlling for VD diagnosis (B = 0.0551, p < 0.001), and path c represents the direct effect of VD diagnosis on T1w/T2w-FLAIR ratio (B = 0.0536, p < 0.05). The indirect effect was significant, indicating partial mediation through vascular risk (a×b = 0.0099, p < 0.05). (B) Bar plot showing the within-group distribution of vascular risk categories (0, 1, 2, ≥3 risk factors) across diagnostic groups (SCI, MCI non-amnestic, MCI amnestic, AD, VD, Other), illustrating that VD patients carry a disproportionately higher vascular risk burden relative to other groups. (C) Bar plot showing mean residualized T1w/T2w-FLAIR ratio across vascular risk categories, with error bars representing standard error, demonstrating a monotonic increase in T1w/T2w-FLAIR ratio with greater vascular risk burden. Vascular risk factors included hypertension, dyslipidemia, diabetes, cerebrovascular disease, peripheral vascular disease, myocardial infarction, congestive heart failure, and angina. All mediation models were adjusted for age, sex, education, site, and diagnosis group. *p < 0.05, **p < 0.01, ***p < 0.001.

## 4. Discussion

We explored T1w/T2w-FLAIR in WMH as a marker of microstructural damage – where lower values are indicative of demyelination. We found that greater age was associated with a greater T1w/T2w-FLAIR ratio, indicative of age-associated vascular damage, and that individuals with VD showed a heightened T1w/T2w-FLAIR ratio, which was partially mediated by cardiovascular burden. Additionally, we found that AD patients showed, on average, a heightened T1w/T2w-FLAIR ratio, and this was partially mediated by plasma Aβ42. Greater amyloid burden (lower plasma Aβ42) was associated with greater demyelination (lower T1w/T2w-FLAIR ratio). These results point to greater heterogeneity of WMH pathology in AD compared with VD, whereby greater amyloid burden (reflected by lower plasma Aβ42) may be associated with demyelination (lower T1w/T2w-FLAIR ratio), while concurrent or alternative vascular mechanisms elevate T1w/T2w-FLAIR ratio, yielding higher average values across the AD group.

Age-related increases in WMH T1w/T2w-FLAIR residuals reflect progressive vascular processes characteristic of cerebral small vessel disease, with greater age associated with greater T1w/T2w-FLAIR ratios independent of clinical diagnosis. As discussed above, positive residuals in older participants are most consistent with a relatively gliotic-cellular response – dense reactive astrogliosis, microglial activation, and increased cellularity – which elevates the lesion T1w signal disproportionately to changes in the FLAIR signal, producing a residual that exceeds the participant’s own NAWM-predicted value (Hase et al., 2018; Silbert et al., 2024; Wardlaw et al., 2019). VD demonstrates the most pronounced T1w/T2w-FLAIR ratio elevations relative to other diagnostic groups, consistent with severe microvascular pathology, including arteriolosclerosis, chronic hypoperfusion, and ischemic injury that may drive tissue damage (Roseborough et al., 2022; Solé-Guardia et al., 2024; Erten-Lyons et al., 2013). While both VD and AD exhibit myelin loss on histopathology, the mechanisms differ fundamentally: in aging and VD, elevated T1w/T2w-FLAIR residuals primarily reflect ischemic-inflammatory injury with reactive gliosis and increased cellularity superimposed on vascular pathology, rather than primary demyelination driven by proteinopathic processes (Ihara et al., 2010; Hase et al., 2018). Postmortem studies show that VD-associated white matter changes are characterized by diffuse rarefaction, loss of oligodendrocytes, gliosis, and arterial wall pathology (Ihara et al., 2010; Erten-Lyons et al., 2013; Kalaria, 2016). The gliotic and inflammatory components of this profile preferentially elevate the lesion T1w signal (through increased cellular density and proteinaceous content) relative to the participant’s own NAWM, yielding positive T1w/T2w-FLAIR residuals despite concurrent myelin damage (Ihara et al., 2010, Hase et al., 2018; Kalaria, 2016). The demyelination observed in VD occurs secondary to hypoxic-ischemic damage and subsequent axonal degeneration, contrasting with primary demyelinating disorders where myelin is the initial target (Ihara et al., 2010). Notably, the VD group showed the highest within-group variability in T1w/T2w-FLAIR residuals despite the highest group mean. While most displayed elevated residuals, some showed much lower ratios that may reflect later-stage lesions in which extensive demyelination and tissue rarefaction begin to dominate over the gliotic-cellular signal. This raises the speculative possibility that residualized T1w/T2w-FLAIR could serve as a staging marker in VD – elevated in earlier, gliotic-ischemic stages and declining in later, rarefactive stages – though longitudinal data would be required to test this directly.

Our finding that vascular comorbidity burden independently predicted higher T1w/T2w-FLAIR ratios, even after accounting for diagnosis and amyloid pathology, supports the interpretation that accumulating vascular risk factors drive microstructural changes characteristic of ischemic-inflammatory injury. The partial mediation of VD diagnosis by vascular burden reinforces that the elevated ratios observed in VD reflect vascular pathophysiology rather than diagnostic categorization alone. This relationship between vascular risk and T1w/T2w-FLAIR ratio elevation provides mechanistic validation for the ischemic-inflammatory model of white matter injury in cerebrovascular disease.

AD patients exhibited elevated mean T1w/T2w-FLAIR ratios, but mediation analyses revealed that this relationship was partially mediated by plasma Aβ42 levels. Lower plasma Aβ42 (reflecting higher brain amyloid burden) was associated with lower T1w/T2w-FLAIR ratios within WMH, suggesting that amyloid-driven demyelination may counteract or dominate overt vascular processes in AD patients with the highest pathological burden. This continuous association is directionally opposite to the group-level effect, in which AD showed higher mean residuals than SCI. We interpret this apparent discrepancy as evidence of within-group heterogeneity in AD. The AD group likely comprises a mixture of individuals: those in whom amyloid-driven demyelination predominates, showing greater amyloid burden and correspondingly lower residuals, and those in whom coexisting small-vessel disease predominates, showing comparatively less amyloid burden and elevated, gliotic residuals. The continuous Aβ42 association captures the former mechanism, whereas the categorical diagnosis effect reflects the average across both. Consistent with this, the AD coefficient increased substantially once plasma Aβ42 was included in the model, whereas the VD coefficient was unchanged, indicating that amyloid burden suppresses the apparent group-level AD effect. This pattern is consistent with evidence that Aβ peptides exert direct cytotoxic effects on oligodendrocytes through multiple mechanisms, including mitochondrial dysfunction, oxidative stress, and apoptosis, impairing myelin maintenance and affecting white matter degradation (Lee et al., 2004; Chen et al., 2006). Recent literature suggests a bidirectional relationship between amyloid and myelin pathology: while Aβ oligomers can directly induce demyelination before axonal degeneration (Maitre et al., 2023; Chen et al., 2006), emerging evidence suggests myelin dysfunction may itself drive amyloid accumulation, creating a pathological feedback loop (Depp et al., 2023; Maitre et al., 2023). Unlike the secondary demyelination observed in VD, where oligodendrocyte loss follows chronic ischemia (Ihara et al., 2010; Hase et al., 2018), AD demonstrates primary proteinopathic injury to oligodendrocytes through direct Aβ toxicity (Lee et al., 2004; Chen et al., 2006;

Chen et al., 2023). The mediation of AD-associated T1w/T2w-FLAIR changes by Aβ42 demonstrates that white matter alterations in AD are potentially mechanistically linked to core proteinopathic processes rather than representing purely secondary vascular consequences, distinguishing AD from VD. We found no association with plasma NfL or GFAP. This could be because these plasma measures reflect relatively diffuse neuroaxonal injury and astrocytic activation (not WMH-local pathology) rather than lesion-local microstructure captured by residualized T1w/T2w-FLAIR (Khalil et al., 2018; Kim et al., 2023; Leipp et al., 2024).

WMH may appear radiologically similar across diagnostic groups but arise from fundamentally different pathological mechanisms that produce opposing effects on tissue microstructure. In VD and aging, elevated T1w/T2w-FLAIR ratios reflect inflammatory and ischemic processes with preserved or reactive tissue responses, where gliosis and increased water content dominate the signal despite concurrent myelin loss. In multiple sclerosis, reduced T1w/T2w ratios indicate active demyelination and tissue loss, where primary destruction of myelin is the dominant pathological process (Boaventura et al., 2022). In AD, both mechanisms may coexist amyloid-driven primary demyelination lowering ratios alongside vascular processes elevating ratios, resulting in heterogeneous pathological profiles within the diagnostic group. This heterogeneity explains why AD shows intermediate average ratios between SCI and VD despite having a distinct mechanistic pathway. The residualized T1w/T2w-FLAIR approach relative to NAWM provides a more nuanced characterization of lesion pathology than volumetric measures alone, potentially enabling identification of distinct pathological endotypes within radiologically homogeneous WMH. Understanding whether individual patients show predominantly demyelinating (low ratio) versus ischemic (high ratio) profiles may inform prognosis.

One application is Amyloid-Related Imaging Abnormalities (ARIA), brain changes visible on MRI in patients receiving anti-amyloid immunotherapy for AD. They appear as either vasogenic edema and/or sulcal effusions (ARIA-edema [ARIA-E]) or as cerebral microhemorrhages and superficial siderosis (ARIA-hemosiderin [ARIA-H]) (Salloway et al., 2022). ARIA risk assessment in anti-amyloid therapy relies on a combination of *APOE* genotype, baseline MRI markers of vascular vulnerability (including cerebral microhemorrhages, superficial siderosis, and overall WMH burden), and treatment-related factors such as dose and titration schedule (Barakos et al., 2022; Cogswell et al., 2022). WMH assessment is largely volumetric or qualitative, serving as a nonspecific proxy for small vessel disease rather than a direct indicator of tissue susceptibility to edema or hemorrhage. The residualized WMH T1w/T2w-FLAIR ratio provides a complementary microstructural measure that may be evaluated as an adjunct marker of white matter vulnerability within existing ARIA risk stratification frameworks.

Specifically, higher ratios may identify lesions with greater vascular permeability, which could lower the threshold for ARIA-E. They may also index small-vessel pathology that co-occurs with hemorrhagic risk in amyloid-laden vasculature. This is consistent with current mechanistic models of ARIA pathophysiology, which emphasize blood-brain barrier dysfunction and vascular amyloid involvement (Barakos et al., 2022; Hampel et al., 2023). Future studies integrating T1w/T2w-FLAIR ratios with established predictors such as *APOE* genotype, baseline microbleed burden, and amyloid load will be required to determine whether this measure provides incremental predictive value for ARIA susceptibility beyond lesion volume alone.

A methodological consideration is that the “T2-weighted” image used in our ratio is in fact FLAIR, and FLAIR signal contains some T1-weighted dependence arising from its inversion-recovery preparation. The T1w/T2w-FLAIR ratio, therefore, captures combined relaxation properties rather than the pure T1/T2 contrast of the original Glasser & Van Essen formulation, and absolute values are not directly comparable to T1w/T2w studies in MS or healthy aging. Our residualization to each participant’s NAWM mitigates but does not eliminate this issue, since both lesion and NAWM signals are subject to this effect. We report this measure under the explicit “T1w/T2w-FLAIR” name to avoid conflation with the pure T1w/T2w literature.

There are several limitations to this study. The cross-sectional design of the dataset prevents any causal inferences about temporal relationships between amyloid accumulation and white matter changes, limiting our ability to determine whether amyloid deposition precedes demyelination or vice versa in the proposed bidirectional relationship. Absence of significant findings in MCI suggests the T1w/T2w-FLAIR ratio may not serve as a preclinical marker, requiring established pathology to manifest detectable changes. This may also reflect a cancellation effect, whereby amyloid-related processes that lower the residualized WMH T1w/T2w-FLAIR ratio and vascular processes that raise it may coexist at this stage, resulting in a net null association rather than an absence of underlying pathology. Limited sample sizes for APOE ε2 and ε4 carriers may have reduced power to detect genetic associations that could clarify inherited vulnerability to specific forms of white matter injury. Missing amyloid PET data limited our ability to fully characterize the relationship between in vivo amyloid burden and WMH microstructure, preventing direct comparison between imaging and plasma markers. The lack of a cognitively normal comparison group (only SCI available) limits interpretation of disease-specific effects versus normal aging trajectories, though this reflects the real-world clinical population where diagnostic uncertainty is common.

Additionally, the sample is primarily Asian, which may limit generalizability to other populations with different genetic backgrounds and environmental exposures. The mediation analyses, while informative, are conducted on a cross-sectional dataset with modest subgroup sample sizes (AD n=125, VD n=88), and the indirect effects should therefore be interpreted as suggestive rather than confirmatory - replication in longitudinal cohorts is required to support causal inference. We also note that diagnostic categorization in this cohort follows current clinical criteria, but most participants likely harbor a mixture of AD-related and vascular pathology to varying degrees. Our findings should accordingly be read as reflecting the predominant pathology of each clinical group rather than mutually exclusive disease categories. Finally, we examined total WMH burden as a single, whole brain measure rather than separating deep, periventricular, and juxtacortical WMHs, which are known to differ in their underlying microvascular and degenerative mechanisms. Future work using anatomically specific ROIs may reveal regionally distinct T1w/T2w-FLAIR signatures. Future mechanistic studies incorporating longitudinal designs, comprehensive biomarker panels including both PET and plasma markers, and postmortem validation are needed to clarify the molecular pathways linking plasma Aβ42 to white matter microstructural integrity and establish temporal precedence in the amyloid-myelin relationship.

The T1w/T2w-FLAIR ratio may have clinical utility in differentiating mixed pathology dementia from pure AD, given VD’s substantially higher ratios and the distinct mechanistic profiles observed across diagnostic groups. VD patients showing markedly elevated ratios likely have predominantly vascular mechanisms, while AD patients with lower ratios may have greater amyloid-driven demyelination. This measure can be derived from standard clinical MRI sequences (T1w and T2w/FLAIR) already acquired in routine dementia evaluations without requiring specialized acquisition protocols. Implementation requires no additional scanning time or specialized sequences, making it broadly accessible for clinical translation across diverse healthcare settings, including those without access to advanced imaging or expensive biomarker assays. The residualized approach could enhance diagnostic precision in cases where vascular and neurodegenerative pathologies overlap, potentially informing treatment decisions by identifying the dominant pathological mechanism driving white matter injury. Our results show that WMH T1w/T2w-FLAIR ratio reveals microstructural heterogeneity in white matter lesions and differentiates VD from AD, which exhibits variable profiles reflecting a mixed amyloid and vascular pathology.

## Data Availability

The data underlying this study were obtained from the Biobank Innovations for Chronic Cerebrovascular Disease with Alzheimer's Disease Study (BICWALZS). BICWALZS data are not publicly available because of participant privacy protections and the terms of the consortium's data governance agreement. De-identified data may be made available to qualified investigators following application to and approval by the BICWALZS consortium. The imaging processing tools used are publicly available at https://github.com/tetra-tools/T1_T2_Ratio and https://github.com/treanus/kul_nis. Analysis code supporting the findings is available from the corresponding author upon reasonable request.

https://github.com/treanus/kul_nis

## Data Availability

https://github.com/treanus/kul_nis

## Acknowledgements

We want to thank the staff of BICWALZS and the Suwon Geriatric Mental Health Centre for their involvement in this study.

## Author contributions

- **Funding acquisition:** SJS, CHH, HWR, BP, JWC, SWS, SHC, SYM, EJK, BCK, YSA, YHC, SH
- **Data collection:** JYC, TD, YP, CA, PCLF, GP, BB, JPFS, DTL, FZL, MSM, ER, FR, CHH, HWR, BP, JWC, SWS, SHC, SYM, EJK, BCK, YSA, YHC, SH, SJS, TKK, TAP
- **Data analysis:** RS, HTK, SJS
- **Drafting:** RS, HTK, SJS
- **Final review of paper:** RS, JYC, TD, YP, CA, PCLF, GP, BB, JPFS, DTL, FZL, MSM, ER, FR, CHH, HWR, BP, JWC, SWS, SHC, SYM, EJK, BCK, YSA, YHC, SH, TKK, TAP, SJS, HTK

## Funding and assistance

This study was conducted using biospecimens and data from the Biobank Innovations for Chronic Cerebrovascular Disease with ALZheimer’s Disease Study (BICWALZS) consortium funded by the Korean National Institute of Health (NIH) research project (Project No. 2024-ER0505-01). This research was supported by grants from the National Research Foundation of Korea (NRF), funded by the Ministry of Science and ICT (RS-2019-NR040055), and grants from the Korea Health Industry Development Institute (KHIDI), funded by the Ministry of Health and Welfare (HR21C1003, HI22C0724, HR22C1734, RS-2023-00267453, RS-2024-00406876 and RS-2025-25455095), to SJS, HWR and CHH. Furthermore, this research was supported by a grant from the Korea Dementia Research Project through the Korea Dementia Research Center (KDRC), funded by the Ministry of Health and Welfare and the Ministry of Science and ICT, Republic of Korea (RS-2024-00339665) to SJS. TAP is supported by the National Institute on Aging (R01AG075336 and R01AG073267). TKK and the Karikari Laboratory were supported by the National Institutes of Health (R01AG083874, U24AG082930, P30AG066468, RF1AG052525, R01AG053952, R37AG023651, RF1AG025516, R01AG073267, R01AG075336, R01AG072641, P01AG025204) and a professorial endowment from the Alzheimer’s Association (24AARFD-1243899).

## Data and resource availability

The datasets generated during and/or analyzed in the current study are available from the corresponding author upon reasonable request.

## Consent Statement

BICWALZS is registered in the Korean National Clinical Trial Registry CRIS (identifier: KCT0003391). Prior to beginning the study, the Institutional Review Board approved the study plan (AJIRB-BMR-SUR-16-362). Written informed consent was obtained from all participants and caregivers. The study was conducted in accordance with the International Conference on Harmonisation guidelines on Good Clinical Practice.

