## Supplementary Material for "Altered T1w/T2w-FLAIR Ratio in White Matter Hyperintensities as an Indicator of Structural Integrity Loss: Association with Alzheimer’s Disease and Vascular Dementia"

**Supplemental Table 1. Association of Residualized T1w/T2w-FLAIR Ratio with Demographics and Clinical Variables (Base Model)**

| **Term** | **B** | **Standard Error** | **P-value** | **CI_lower_95** | **CI_upper_95** | **B (Imputed)** | **P-value (Imputed)** |
| --- | --- | --- | --- | --- | --- | --- | --- |
| **Intercept** | **-0.364** | **0.066** | **<0.001** | **-0.493** | **-0.235** | **-0.340** | **<0.001** |
| **Age** | **0.005** | **0.0009** | **<0.001** | **0.003** | **0.006** | **0.004** | **<0.001** |
| Sex (Male) | 0.015 | 0.0146 | 0.31 | -0.014 | 0.043 | 0.017 | 0.25 |
| Education (years) | 0.0003 | 0.0015 | 0.84 | -0.0027 | 0.0033 | -0.0004 | 0.82 |
| Days Between Scan and Visit | -0.0004 | 0.0003 | 0.12 | -0.0010 | 0.0001 | -0.0003 | 0.30 |
| Site: Chonnam | 0.028 | 0.0385 | 0.47 | -0.048 | 0.104 | 0.027 | 0.49 |
| Site: Gwangju | 0.064 | 0.0399 | 0.11 | -0.014 | 0.143 | 0.049 | 0.29 |
| **Site: Inha** | **0.039** | **0.0189** | **0.04** | **0.002** | **0.076** | **0.039** | **0.049** |
| Site: Pusan | 0.020 | 0.0328 | 0.54 | -0.044 | 0.084 | 0.025 | 0.47 |
| **Site: Samsung** | **0.142** | **0.0219** | **<0.001** | **0.099** | **0.185** | **0.134** | **<0.001** |
| Site: Suwon | 0.017 | 0.0183 | 0.35 | -0.019 | 0.053 | 0.014 | 0.42 |

**Notes:** Model adjusted for age, sex, education, site, and days between scan and visit. Reference categories:
Sex = Female; Site = Ajou. Multiple imputations by chained equations (MICE) were performed using the random forest method with m = 5 imputations. Results presented for both the complete-case analysis (Base Model) and the imputed datasets. Linear regression with residualized T1w/T2w-FLAIR ratio as the dependent variable. CI = Confidence Interval (95%). Bold values indicate statistically significant associations (p < 0.05) in the base model.

**Supplemental Table 2. Association of Residualized T1w/T2w-FLAIR Ratio with Demographics and Clinical group**

| **Term** | **B** | **Standard Error** | **P-value** | **CI_lower_95** | **CI_upper_95** | **B (Imputed)** | **P-value (Imputed)** |
| --- | --- | --- | --- | --- | --- | --- | --- |
| **Intercept** | **-0.374** | **0.067** | **<0.001** | **-0.506** | **-0.242** | **-0.351** | **<0.001** |
| **Age** | **0.004** | **0.001** | **<0.001** | **0.002** | **0.006** | **0.004** | **<0.001** |
| Sex (Male) | 0.01 | 0.015 | 0.52 | -0.019 | 0.038 | 0.012 | 0.41 |
| Education (years) | 0.0004 | 0.002 | 0.79 | -0.003 | 0.003 | -0.0002 | 0.89 |
| Days Between Scan and Visit | -0.0004 | 0.0003 | 0.14 | -0.001 | 0.0001 | -0.0003 | 0.35 |
| Site: Chonnam | 0.016 | 0.039 | 0.67 | -0.06 | 0.093 | 0.016 | 0.69 |
| *Site: Gwangju* | *0.086* | *0.041* | *0.038* | *0.005* | *0.167* | *0.069* | *0.16* |
| **Site: Inha** | **0.045** | **0.019** | **0.017** | **0.008** | **0.083** | **0.045** | **0.026** |
| Site: Pusan | 0.02 | 0.033 | 0.55 | -0.045 | 0.084 | 0.025 | 0.48 |
| **Site: Samsung** | **0.138** | **0.022** | **<0.001** | **0.095** | **0.181** | **0.131** | **<0.001** |
| Site: Suwon | 0.023 | 0.019 | 0.23 | -0.014 | 0.06 | 0.02 | 0.26 |
| Group: MCI non-amnestic | -0.001 | 0.027 | 0.98 | -0.054 | 0.053 | -0.005 | 0.84 |
| Group: MCI amnestic | 0.025 | 0.023 | 0.28 | -0.02 | 0.07 | 0.022 | 0.32 |
| **Group: AD** | **0.052** | **0.025** | **0.038** | **0.003** | **0.101** | **0.049** | **0.044** |
| **Group: VD** | **0.063** | **0.026** | **0.016** | **0.012** | **0.115** | **0.06** | **0.019** |
| Group: Other | 0.034 | 0.039 | 0.38 | -0.043 | 0.11 | 0.025 | 0.52 |

**Abbreviations:** AD, Alzheimer's disease; MCI, mild cognitive impairment; SCI, subjective cognitive impairment; VD, vascular dementia; CI, confidence interval.

**Notes:** Model adjusted for age, sex, education, site, and days between scan and visit. Dependent variable is residualized T1w/T2w-FLAIR ratio (global ratio adjusted for normal-appearing white matter). Reference categories: Sex = Female; Site = Ajou; Group = SCI. Multiple imputations by chained equations (MICE) performed using random forest method with m = 5 imputations. Results presented for both complete case analysis and imputed datasets. Linear regression with residualized T1w/T2w-FLAIR ratio as the dependent variable. CI = 95% confidence interval. Italicized values indicate associations that were statistically significant in only one of the two analyses (either complete-case or imputed analysis, but not both).

**Supplemental Table 3. Association of Residualized T1w/T2w-FLAIR Ratio with Plasma Aβ42**

| **Variable** | **B** | **Standard Error** | **P-value** | **B (Imputed)** | **P-value (Imputed)** |
| --- | --- | --- | --- | --- | --- |
| **Intercept** | **-0.400** | **0.077** | **<0.001** | **-0.361** | **<0.001** |
| **Age** | **0.004** | **0.001** | **<0.001** | **0.004** | **<0.001** |
| Sex (Male) | 0.004 | 0.016 | 0.78 | 0.014 | 0.33 |
| Education (years) | 0.001 | 0.002 | 0.71 | 0.0002 | 0.89 |
| Site: Chonnam | 0.011 | 0.042 | 0.8 | 0.017 | 0.67 |
| Site: Gwangju | 0.105 | 0.046 | 0.024 | 0.087 | 0.041 |
| Site: Inha | 0.045 | 0.022 | 0.045 | 0.05 | 0.005 |
| Site: Pusan | -0.007 | 0.034 | 0.85 | 0.008 | 0.81 |
| **Site: Samsung** | **0.114** | **0.024** | **<0.001** | **0.129** | **<0.001** |
| Site: Suwon | 0.0003 | 0.02 | 0.99 | 0.02 | 0.24 |
| Group: MCI non-amnestic | 0.004 | 0.03 | 0.89 | 0.007 | 0.78 |
| Group: MCI amnestic | 0.030 | 0.026 | 0.24 | 0.027 | 0.22 |
| **Group: AD** | **0.071** | **0.029** | **0.013** | **0.059** | **0.013** |
| **Group: VD** | **0.060** | **0.029** | **0.043** | **0.062** | **0.011** |
| Group: Other | 0.034 | 0.043 | 0.43 | 0.036 | 0.33 |
| **Plasma Aβ42** | **0.010** | **0.004** | **0.009** | **0.009** | **0.023** |
| Days Between Scan and Visit | -0.0005 | 0.0003 | 0.09 | -0.0004 | 0.22 |

Abbreviations: AD, Alzheimer's disease; Aβ, amyloid-beta; CI, confidence interval; MCI, mild cognitive impairment; SCI, subjective cognitive impairment; VD, vascular dementia.

Notes: Model adjusted for age, sex, education, site, cognitive diagnosis, and days between scan and visit. The dependent variable is residualized T1w/T2w-FLAIR ratio (global ratio adjusted for normal-appearing white matter). Plasma Aβ42 was modeled as a continuous predictor. Reference categories: Group = SCI; Sex = Female; Site = Ajou. Multiple imputation by chained equations (MICE) was performed using the random forest method with m = 5 imputations. Results are presented for both the complete-case analysis and the pooled imputed datasets. All estimates represent unstandardized regression coefficients (B). Italicized values indicate associations that were statistically significant in only one of the two analyses (either complete-case or imputed, but not both).

**Supplemental Table 4. Association of Residualized T1w/T2w-FLAIR Ratio with Plasma Aβ40**

| **Term** | **B** | **Standard Error** | **P-value** | **CI_lower_95** | **CI_upper_95** | **B (Imputed)** | **P-value (Imputed)** |
| --- | --- | --- | --- | --- | --- | --- | --- |
| **Intercept** | **-0.406** | **0.075** | **<0.001** | **-0.554** | **-0.259** | **-0.355** | **<0.001** |
| **Age** | **0.004** | **0.001** | **<0.001** | **0.002** | **0.006** | **0.004** | **<0.001** |
| Sex (Male) | -0.001 | 0.016 | 0.97 | -0.031 | 0.03 | 0.012 | 0.42 |
| Education (years) | 0.001 | 0.002 | 0.56 | -0.002 | 0.004 | -0.0004 | 0.82 |
| Days Between Scan and Visit | -0.0004 | 0.0003 | 0.14 | -0.001 | 0.0001 | -0.0003 | 0.38 |
| Site: Chonnam | 0.015 | 0.04 | 0.71 | -0.063 | 0.092 | 0.019 | 0.63 |
| *Site: Gwangju* | *0.108* | *0.044* | *0.014* | *0.022* | *0.195* | *0.078* | *0.12* |
| **Site: Inha** | **0.053** | **0.021** | **0.013** | **0.011** | **0.094** | **0.049** | **0.016** |
| Site: Pusan | -0.001 | 0.034 | 0.99 | -0.067 | 0.066 | 0.012 | 0.74 |
| **Site: Samsung** | **0.127** | **0.023** | **<0.001** | **0.082** | **0.173** | **0.126** | **<0.001** |
| Site: Suwon | 0.006 | 0.02 | 0.77 | -0.033 | 0.045 | 0.011 | 0.56 |
| Group: MCI non-amnestic | 0.015 | 0.029 | 0.62 | -0.043 | 0.072 | -0.001 | 0.96 |
| Group: MCI amnestic | 0.038 | 0.025 | 0.13 | -0.011 | 0.088 | 0.022 | 0.32 |
| **Group: AD** | **0.073** | **0.027** | **0.008** | **0.02** | **0.127** | **0.053** | **0.028** |
| **Group: VD** | **0.074** | **0.029** | **0.01** | **0.018** | **0.13** | **0.059** | **0.022** |
| Group: Other | 0.054 | 0.041 | 0.19 | -0.027 | 0.136 | 0.03 | 0.42 |
| **Plasma Aβ40** | **0.0006** | **0.0002** | **0.005** | **0.0002** | **0.001** | **0.0005** | **0.02** |

**Abbreviations:** AD, Alzheimer's disease; Aβ40, amyloid-beta 40; MCI, mild cognitive impairment; SCI, subjective cognitive impairment; VD, vascular dementia; CI, confidence interval.

**Notes:** Model adjusted for age, sex, education, site, cognitive diagnosis, and days between scan and visit. The dependent variable is residualized T1w/T2w-FLAIR ratio (global ratio adjusted for normal-appearing white matter). Plasma Aβ40 is modeled as a continuous variable. Reference categories: Group = SCI; Sex = Female; Site = Ajou. Multiple imputations by chained equations (MICE) were performed using random forest method with m = 5 imputations. Results presented for both the complete-case analysis and the imputed datasets. Linear regression with residualized T1w/T2w-FLAIR ratio as the dependent variable. CI = 95% confidence interval. All estimates represent unstandardized regression coefficients (B). Italicized values indicate associations that were statistically significant in only one of the two analyses (either complete-case or imputed analysis, but not both).

**Supplemental Table 5. Association of Residualized T1w/T2w-FLAIR Ratio with APOE Genotype**

| **Term** | **B** | **Standard Error** | **P-value** | **CI_lower_95** | **CI_upper_95** | **B (Imputed)** | **P-value (Imputed)** |
| --- | --- | --- | --- | --- | --- | --- | --- |
| **Intercept** | **-0.378** | **0.069** | **<0.001** | **-0.513** | **-0.242** | **-0.356** | **<0.001** |
| **Age** | **0.004** | **0.001** | **<0.001** | **0.003** | **0.006** | **0.004** | **<0.001** |
| Sex (Male) | 0.009 | 0.015 | 0.54 | -0.02 | 0.038 | 0.012 | 0.42 |
| Education (years) | 0.0004 | 0.002 | 0.78 | -0.003 | 0.003 | -0.0002 | 0.89 |
| Days Between Scan and Visit | -0.0004 | 0.0003 | 0.14 | -0.001 | 0.0001 | -0.0003 | 0.33 |
| Site: Chonnam | 0.019 | 0.039 | 0.62 | -0.057 | 0.096 | 0.018 | 0.65 |
| *Site: Gwangju* | *0.087* | *0.041* | *0.036* | *0.006* | *0.168* | *0.07* | *0.16* |
| **Site: Inha** | **0.044** | **0.019** | **0.021** | **0.007** | **0.081** | **0.043** | **0.03** |
| Site: Pusan | 0.018 | 0.033 | 0.59 | -0.047 | 0.083 | 0.023 | 0.5 |
| **Site: Samsung** | **0.137** | **0.022** | **<0.001** | **0.094** | **0.181** | **0.13** | **<0.001** |
| Site: Suwon | 0.023 | 0.019 | 0.22 | -0.014 | 0.06 | 0.02 | 0.25 |
| Group: MCI non-amnestic | -0.003 | 0.027 | 0.91 | -0.057 | 0.051 | -0.007 | 0.78 |
| Group: MCI amnestic | 0.022 | 0.023 | 0.34 | -0.023 | 0.068 | 0.02 | 0.38 |
| Group: AD | 0.046 | 0.026 | 0.075 | -0.005 | 0.096 | 0.043 | 0.081 |
| **Group: VD** | **0.062** | **0.026** | **0.019** | **0.01** | **0.113** | **0.059** | **0.022** |
| Group: Other | 0.031 | 0.039 | 0.43 | -0.046 | 0.107 | 0.022 | 0.56 |
| APOE: E3 carrier | -0.006 | 0.021 | 0.78 | -0.047 | 0.035 | -0.001 | 0.95 |
| APOE: E4 carrier | 0.01 | 0.023 | 0.66 | -0.035 | 0.055 | 0.013 | 0.55 |

**Abbreviations:** AD, Alzheimer's disease; APOE, apolipoprotein E; MCI, mild cognitive impairment; SCI, subjective cognitive impairment; VD, vascular dementia; CI, confidence interval.

**Notes:** Model adjusted for age, sex, education, site, cognitive diagnosis, and days between scan and visit. The dependent variable is residualized T1w/T2w-FLAIR ratio (global ratio adjusted for normal-appearing white matter). Reference categories: Sex = Female; Site = Ajou; APOE = E2_carrier (e22, e23), Group: SCI. Multiple imputations by chained equations (MICE) were performed using random forest method with m = 5 imputations. Results presented for both the complete-case analysis and the imputed datasets. Linear regression with residualized T1w/T2w-FLAIR ratio as the dependent variable. CI = 95% confidence interval. All estimates represent unstandardized regression coefficients (B). Italicized values indicate associations that were statistically significant in only one of the two analyses (either complete-case or imputed analysis, but not both).

**Supplemental Table 6. Association of Residualized T1w/T2w-FLAIR Ratio with Amyloid PET Status**

| **Term** | **B** | **Standard Error** | **P-value** | **CI_lower_95** | **CI_upper_95** | **B (Imputed)** | **P-value (Imputed)** |
| --- | --- | --- | --- | --- | --- | --- | --- |
| **Intercept** | **-0.357** | **0.073** | **<0.001** | **-0.5** | **-0.214** | **-0.354** | **<0.001** |
| **Age** | **0.004** | **0.001** | **<0.001** | **0.002** | **0.006** | **0.004** | **<0.001** |
| Sex (Male) | 0.012 | 0.016 | 0.45 | -0.019 | 0.043 | 0.012 | 0.43 |
| Education (years) | 0.001 | 0.002 | 0.57 | -0.002 | 0.004 | -0.0002 | 0.91 |
| Days Between Scan and Visit | -0.0003 | 0.0003 | 0.28 | -0.0009 | 0.0003 | -0.0003 | 0.35 |
| Site: Chonnam | -0.002 | 0.042 | 0.97 | -0.083 | 0.08 | 0.014 | 0.71 |
| Site: Gwangju | 0.056 | 0.044 | 0.2 | -0.03 | 0.142 | 0.068 | 0.16 |
| *Site: Inha* | *0.032* | *0.021* | *0.13* | *-0.009* | *0.072* | *0.045* | *0.028* |
| Site: Pusan | 0.006 | 0.034 | 0.85 | -0.06 | 0.073 | 0.026 | 0.48 |
| **Site: Samsung** | **0.125** | **0.024** | **<0.001** | **0.078** | **0.172** | **0.131** | **<0.001** |
| Site: Suwon | 0.035 | 0.021 | 0.092 | -0.006 | 0.076 | 0.019 | 0.27 |
| Group: MCI non-amnestic | -0.022 | 0.03 | 0.45 | -0.081 | 0.036 | -0.005 | 0.86 |
| Group: MCI amnestic | 0.032 | 0.026 | 0.23 | -0.02 | 0.083 | 0.023 | 0.3 |
| *Group: AD* | *0.04* | *0.03* | *0.18* | *-0.019* | *0.099* | *0.053* | *0.04* |
| *Group: VD* | *0.045* | *0.029* | *0.12* | *-0.011* | *0.102* | *0.06* | *0.02* |
| Group: Other | -0.002 | 0.042 | 0.97 | -0.083 | 0.08 | 0.025 | 0.51 |
| Amyloid Positive | -0.016 | 0.018 | 0.36 | -0.051 | 0.018 | -0.006 | 0.75 |

**Abbreviations:** AD, Alzheimer's disease; MCI, mild cognitive impairment; PET, positron emission tomography; SCI, subjective cognitive impairment; VD, vascular dementia; CI, confidence interval.

**Notes:** Model adjusted for age, sex, education, site, cognitive diagnosis, and days between scan and visit. The dependent variable is residualized T1w/T2w-FLAIR ratio (global ratio adjusted for normal-appearing white matter). Reference categories: Amyloid PET status = negative; Group = SCI; Sex = Female; Site = Ajou. Multiple imputations by chained equations (MICE) were performed using random forest method with m = 5 imputations. Results presented for both the complete-case analysis and the imputed datasets. Linear regression with residualized T1w/T2w-FLAIR ratio as the dependent variable. CI = 95% confidence interval. All estimates represent unstandardized regression coefficients (B). Italicized values indicate associations that were statistically significant in only one of the two analyses (either complete-case or imputed analysis, but not both).

**Supplemental Table 7. Association of Residualized T1w/T2w-FLAIR Ratio with Plasma p-tau217**

| **Term** | **B** | **Standard Error** | **P-value** | **CI_lower_95** | **CI_upper_95** | **B (Imputed)** | **P-value (Imputed)** |
| --- | --- | --- | --- | --- | --- | --- | --- |
| **Intercept** | **-0.363** | **0.067** | **<0.001** | **-0.495** | **-0.231** | **-0.343** | **<0.001** |
| **Age** | **0.004** | **0.001** | **<0.001** | **0.002** | **0.006** | **0.004** | **<0.001** |
| Sex (Male) | 0.01 | 0.015 | 0.48 | -0.018 | 0.039 | 0.013 | 0.38 |
| Education (years) | 0.0003 | 0.002 | 0.86 | -0.003 | 0.003 | -0.0003 | 0.83 |
| Days Between Scan and Visit | -0.0004 | 0.0003 | 0.15 | -0.001 | 0.0001 | -0.0003 | 0.35 |
| Site: Chonnam | 0.02 | 0.039 | 0.61 | -0.056 | 0.096 | 0.019 | 0.63 |
| *Site: Gwangju* | *0.086* | *0.041* | *0.038* | *0.005* | *0.167* | *0.069* | *0.16* |
| *Site: Inha* | *0.044* | *0.019* | *0.022* | *0.006* | *0.081* | *0.043* | *0.033* |
| Site: Pusan | 0.015 | 0.033 | 0.64 | -0.049 | 0.08 | 0.021 | 0.55 |
| **Site: Samsung** | **0.137** | **0.022** | **<0.001** | **0.094** | **0.18** | **0.129** | **<0.001** |
| Site: Suwon | 0.023 | 0.019 | 0.22 | -0.014 | 0.06 | 0.02 | 0.25 |
| Group: MCI non-amnestic | -0.002 | 0.027 | 0.95 | -0.055 | 0.052 | -0.006 | 0.82 |
| Group: MCI amnestic | 0.021 | 0.023 | 0.37 | -0.025 | 0.066 | 0.018 | 0.41 |
| Group: AD | 0.038 | 0.027 | 0.15 | -0.014 | 0.09 | 0.037 | 0.15 |
| **Group: VD** | **0.062** | **0.026** | **0.019** | **0.01** | **0.113** | **0.058** | **0.023** |
| Group: Other | 0.033 | 0.039 | 0.4 | -0.044 | 0.109 | 0.024 | 0.53 |
| Plasma pTau217 | 0.079 | 0.055 | 0.15 | -0.03 | 0.188 | 0.07 | 0.17 |

**Abbreviations:** AD, Alzheimer's disease; MCI, mild cognitive impairment; p-tau217, phosphorylated tau217; SCI, subjective cognitive impairment; VD, vascular dementia; CI, confidence interval.

**Notes:** Model adjusted for age, sex, education, site, cognitive diagnosis, and days between scan and visit. The dependent variable is residualized T1w/T2w-FLAIR ratio (global ratio adjusted for normal-appearing white matter). Reference categories: Group = SCI; Sex = Female; Site = Ajou. Multiple imputations by chained equations (MICE) were performed using random forest method with m = 5 imputations. Results presented for both the complete-case analysis and the imputed datasets. Linear regression with residualized T1w/T2w-FLAIR ratio as the dependent variable. CI = 95% confidence interval. All estimates represent unstandardized regression coefficients (B). Italicized values indicate associations that were statistically significant in only one of the two analyses (either complete-case or imputed analysis, but not both).

**Supplemental Table 8. Association of Residualized T1w/T2w-FLAIR Ratio with Plasma NfL**

| **Term** | **B** | **Standard Error** | **P-value** | **CI_lower_95** | **CI_upper_95** | **B (Imputed)** | | **P-value (Imputed)** |
| --- | --- | --- | --- | --- | --- | --- | --- | --- |
| **Intercept** | **-0.402** | **0.071** | **<0.001** | **-0.541** | **-0.262** | **-0.341** | **<0.001** | |
| **Age** | **0.004** | **0.001** | **<0.001** | **0.003** | **0.006** | **0.004** | **<0.001** | |
| Sex (Male) | 0.002 | 0.015 | 0.92 | -0.028 | 0.032 | 0.01 | 0.49 | |
| Education (years) | 0.001 | 0.002 | 0.54 | -0.002 | 0.004 | -0.0001 | 0.93 | |
| Days Between Scan and Visit | -0.0004 | 0.0003 | 0.12 | -0.001 | 0.0001 | -0.0003 | 0.34 | |
| Site: Chonnam | 0.015 | 0.039 | 0.7 | -0.062 | 0.092 | 0.016 | 0.68 | |
| *Site: Gwangju* | *0.096* | *0.042* | *0.023* | *0.013* | *0.179* | *0.071* | *0.15* | |
| **Site: Inha** | **0.05** | **0.02** | **0.015** | **0.01** | **0.09** | **0.043** | **0.034** | |
| Site: Pusan | 0.021 | 0.033 | 0.54 | -0.045 | 0.086 | 0.026 | 0.45 | |
| **Site: Samsung** | **0.138** | **0.022** | **<0.001** | **0.094** | **0.182** | **0.131** | **<0.001** | |
| Site: Suwon | 0.022 | 0.019 | 0.24 | -0.015 | 0.06 | 0.022 | 0.22 | |
| Group: MCI non-amnestic | 0.001 | 0.028 | 0.97 | -0.054 | 0.056 | -0.005 | 0.86 | |
| Group: MCI amnestic | 0.03 | 0.024 | 0.21 | -0.017 | 0.078 | 0.022 | 0.33 | |
| *Group: AD* | *0.054* | *0.026* | *0.038* | *0.003* | *0.106* | *0.047* | *0.055* | |
| **Group: VD** | **0.068** | **0.028** | **0.014** | **0.014** | **0.122** | **0.059** | **0.023** | |
| Group: Other | 0.036 | 0.04 | 0.37 | -0.042 | 0.114 | 0.022 | 0.57 | |
| Plasma NFL | 0.0003 | 0.0002 | 0.25 | -0.0002 | 0.0007 | 0.0002 | 0.3 | |

**Abbreviations:** AD, Alzheimer's disease; MCI, mild cognitive impairment; NfL, neurofilament light chain; SCI, subjective cognitive impairment; VD, vascular dementia; CI, confidence interval.

**Notes:** Model adjusted for age, sex, education, site, cognitive diagnosis, and days between scan and visit. The dependent variable is residualized T1w/T2w-FLAIR ratio (global ratio adjusted for normal-appearing white matter). Plasma NfL is modeled as a continuous variable. Reference categories: Group = SCI; Sex = Female; Site = Ajou. Multiple imputations by chained equations (MICE) were performed using random forest method with m = 5 imputations. Results presented for both the complete-case analysis and the imputed datasets. Linear regression with residualized T1w/T2w-FLAIR ratio as the dependent variable. CI = 95% confidence interval. All estimates represent unstandardized regression coefficients (B). Italicized values indicate associations that were statistically significant in only one of the two analyses (either complete-case or imputed analysis, but not both).

**Supplemental Table 9. Association of Residualized T1w/T2w-FLAIR Ratio with Plasma GFAP**

| **Term** | **B** | **Standard Error** | **P-value** | **CI_lower_95** | **CI_upper_95** | **B (Imputed)** | **P-value (Imputed)** |
| --- | --- | --- | --- | --- | --- | --- | --- |
| **Intercept** | **-0.416** | **0.07** | **<0.001** | **-0.554** | **-0.278** | **-0.351** | **<0.001** |
| **Age** | **0.005** | **0.001** | **<0.001** | **0.003** | **0.007** | **0.004** | **<0.001** |
| Sex (Male) | 0.003 | 0.015 | 0.83 | -0.027 | 0.033 | 0.012 | 0.41 |
| Education (years) | 0.001 | 0.002 | 0.57 | -0.002 | 0.004 | -0.0002 | 0.89 |
| Days Between Scan and Visit | -0.0004 | 0.0003 | 0.12 | -0.001 | 0.0001 | -0.0003 | 0.35 |
| Site: Chonnam | 0.014 | 0.039 | 0.72 | -0.063 | 0.091 | 0.016 | 0.69 |
| *Site: Gwangju* | *0.093* | *0.042* | *0.028* | *0.01* | *0.176* | *0.069* | *0.16* |
| **Site: Inha** | **0.052** | **0.02** | **0.011** | **0.012** | **0.092** | **0.045** | **0.027** |
| Site: Pusan | 0.018 | 0.033 | 0.58 | -0.047 | 0.083 | 0.025 | 0.48 |
| **Site: Samsung** | **0.138** | **0.023** | **<0.001** | **0.093** | **0.182** | **0.131** | **<0.001** |
| Site: Suwon | 0.02 | 0.019 | 0.3 | -0.018 | 0.057 | 0.02 | 0.26 |
| Group: MCI non-amnestic | 0.001 | 0.028 | 0.97 | -0.054 | 0.057 | -0.005 | 0.84 |
| Group: MCI amnestic | 0.032 | 0.024 | 0.19 | -0.016 | 0.079 | 0.022 | 0.33 |
| *Group: AD* | *0.059* | *0.027* | *0.026* | *0.007* | *0.112* | *0.049* | *0.052* |
| **Group: VD** | **0.071** | **0.028** | **0.01** | **0.017** | **0.126** | **0.06** | **0.02** |
| Group: Other | 0.041 | 0.04 | 0.31 | -0.037 | 0.119 | 0.024 | 0.52 |
| Plasma GFAP | -0.00002 | 0.00006 | 0.76 | -0.00013 | 0.00009 | 0 | 0.93 |

**Abbreviations:** AD, Alzheimer's disease; GFAP, glial fibrillary acidic protein; MCI, mild cognitive impairment; SCI, subjective cognitive impairment; VD, vascular dementia.

**Notes:** Model adjusted for age, sex, education, site, cognitive diagnosis, and days between scan and visit. The dependent variable is residualized T1w/T2w-FLAIR ratio (global ratio adjusted for normal-appearing white matter). Plasma GFAP is modeled as a continuous variable. Reference categories: Group = SCI; Sex = Female; Site = Ajou. Multiple imputations by chained equations (MICE) were performed using random forest method with m = 5 imputations. Results presented for both the complete-case analysis and the imputed datasets. Linear regression with residualized T1w/T2w-FLAIR ratio as the dependent variable. CI = 95% confidence interval. All estimates represent unstandardized regression coefficients (B). Italicized values indicate associations that were statistically significant in only one of the two analyses (either complete-case or imputed analysis, but not both).

**Supplemental Table 10. Association of Residualized T1w/T2w-FLAIR Ratio with Plasma Aβ42/40 Ratio**

| **Term** | **B** | **Standard Error** | **P-value** | **CI_lower_95** | **CI_upper_95** | **B (Imputed)** | **P-value (Imputed)** |
| --- | --- | --- | --- | --- | --- | --- | --- |
| **Intercept** | **-0.401** | **0.078** | **<0.001** | **-0.555** | **-0.247** | **-0.352** | **<0.001** |
| **Age** | **0.005** | **0.001** | **<0.001** | **0.003** | **0.007** | **0.004** | **<0.001** |
| Sex (Male) | 0.004 | 0.016 | 0.81 | -0.028 | 0.036 | 0.012 | 0.42 |
| Education (years) | 0.001 | 0.002 | 0.65 | -0.003 | 0.004 | -0.0002 | 0.91 |
| Days Between Scan and Visit | -0.0006 | 0.0003 | 0.08 | -0.0012 | 0.0001 | -0.0003 | 0.35 |
| Site: Chonnam | 0.009 | 0.043 | 0.83 | -0.075 | 0.093 | 0.015 | 0.7 |
| *Site: Gwangju* | *0.096* | *0.047* | *0.04* | *0.004* | *0.188* | *0.069* | *0.16* |
| **Site: Inha** | **0.041** | **0.022** | **0.067** | **-0.003** | **0.085** | **0.045** | **0.026** |
| Site: Pusan | 0.009 | 0.034 | 0.8 | -0.059 | 0.076 | 0.025 | 0.48 |
| **Site: Samsung** | **0.12** | **0.024** | **<0.001** | **0.072** | **0.168** | **0.131** | **<0.001** |
| Site: Suwon | 0.011 | 0.02 | 0.58 | -0.029 | 0.051 | 0.02 | 0.25 |
| Group: MCI non-amnestic | -0.001 | 0.03 | 0.98 | -0.06 | 0.059 | -0.005 | 0.84 |
| Group: MCI amnestic | 0.031 | 0.026 | 0.24 | -0.021 | 0.083 | 0.022 | 0.32 |
| **Group: AD** | **0.069** | **0.029** | **0.018** | **0.012** | **0.126** | **0.05** | **0.043** |
| **Group: VD** | **0.064** | **0.03** | **0.034** | **0.005** | **0.123** | **0.06** | **0.019** |
| Group: Other | 0.028 | 0.044 | 0.52 | -0.058 | 0.115 | 0.024 | 0.53 |
| Aβ42/40 Ratio (normalized) | 0.009 | 0.007 | 0.21 | -0.005 | 0.023 | 0.002 | 0.76 |

**Abbreviations:** AD, Alzheimer’s disease; Aβ, amyloid-beta; CI, confidence interval; MCI, mild cognitive impairment; SCI, subjective cognitive impairment; VD, vascular dementia.

**Notes:** Model adjusted for age, sex, education, site, cognitive diagnosis, and days between scan and visit. The dependent variable is residualized T1w/T2w-FLAIR ratio (global ratio adjusted for normal-appearing white matter). Plasma Aβ42/40 ratio (normalized) was modeled as a continuous predictor. Reference categories: Group = SCI; Sex = Female; Site = Ajou. Multiple imputations by chained equations (MICE) were performed using random forest method with m = 5 imputations. Results presented for both the complete-case analysis and the imputed datasets. Linear regression with residualized T1w/T2w-FLAIR ratio as the dependent variable. CI = 95% confidence interval. All estimates represent unstandardized regression coefficients (B). Italicized values indicate associations that were statistically significant in only one of the two analyses (either complete-case or imputed analysis, but not both).

**Supplemental Table 11. Association of Residualized T1w/T2w-FLAIR Ratio with Plasma pTau217/Aβ42 Ratio**

| **Term** | **B** | **Standard Error** | **P-value** | **CI_lower_95** | **CI_upper_95** | **B (Imputed)** | **P-value (Imputed)** |
| --- | --- | --- | --- | --- | --- | --- | --- |
| **Intercept** | **-0.385** | **0.078** | **<0.001** | **-0.539** | **-0.232** | **-0.344** | **<0.001** |
| **Age** | **0.005** | **0.001** | **<0.001** | **0.003** | **0.006** | **0.004** | **<0.001** |
| Sex (Male) | 0.006 | 0.016 | 0.73 | -0.026 | 0.037 | 0.012 | 0.41 |
| Education (years) | 0.001 | 0.002 | 0.75 | -0.003 | 0.004 | -0.0003 | 0.85 |
| Days Between Scan and Visit | -0.0006 | 0.0003 | 0.069 | -0.0012 | 0 | -0.0003 | 0.33 |
| Site: Chonnam | 0.011 | 0.043 | 0.79 | -0.072 | 0.095 | 0.016 | 0.67 |
| Site: Gwangju | 0.091 | 0.047 | 0.051 | 0 | 0.183 | 0.067 | 0.18 |
| *Site: Inha* | *0.038* | *0.023* | *0.096* | *-0.007* | *0.082* | *0.043* | *0.038* |
| Site: Pusan | 0.007 | 0.034 | 0.84 | -0.06 | 0.074 | 0.025 | 0.48 |
| **Site: Samsung** | **0.119** | **0.024** | **<0.001** | **0.071** | **0.167** | **0.131** | **<0.001** |
| Site: Suwon | 0.009 | 0.02 | 0.67 | -0.031 | 0.048 | 0.021 | 0.24 |
| Group: MCI non-amnestic | -0.004 | 0.03 | 0.91 | -0.062 | 0.055 | -0.006 | 0.81 |
| Group: MCI amnestic | 0.026 | 0.026 | 0.31 | -0.025 | 0.078 | 0.02 | 0.37 |
| *Group: AD* | *0.06* | *0.03* | *0.048* | *0* | *0.12* | *0.044* | *0.086* |
| **Group: VD** | **0.058** | **0.03** | **0.0496** | **0** | **0.117** | **0.059** | **0.022** |
| Group: Other | 0.028 | 0.044 | 0.52 | -0.058 | 0.114 | 0.023 | 0.54 |
| pTau217/Aβ42 Ratio (normalized) | 0.001 | 0.008 | 0.9 | -0.015 | 0.017 | 0.005 | 0.49 |

**Abbreviations:** AD, Alzheimer’s disease; Aβ, amyloid-beta; p-tau217, phosphorylated tau217; CI, confidence interval; MCI, mild cognitive impairment; SCI, subjective cognitive impairment; VD, vascular dementia.

**Notes:** Model adjusted for age, sex, education, site, cognitive diagnosis, and days between scan and visit. The dependent variable is residualized T1w/T2w-FLAIR ratio (global ratio adjusted for normal-appearing white matter). Plasma pTau217/Aβ42 ratio (normalized) was modeled as a continuous predictor. Reference categories: Group = SCI; Sex = Female; Site = Ajou. Multiple imputations by chained equations (MICE) were performed using random forest method with m = 5 imputations. Results presented for both the complete-case analysis and the imputed datasets. Linear regression with residualized T1w/T2w-FLAIR ratio as the dependent variable. CI = 95% confidence interval. All estimates represent unstandardized regression coefficients (B). Italicized values indicate associations that were statistically significant in only one of the two analyses (either complete-case or imputed analysis, but not both).

**Supplemental Table 12. Mediation Analysis Results**

| **Model** | **Path** | **Effect** | **Coefficient (B)** | **Standard Error** | **P-value** |
| --- | --- | --- | --- | --- | --- |
| **Model 1: VD Status → Aβ40→ T1w/T2w-FLAIR Ratio** | a (VD → Aβ40) | Direct | 0.539 | 5.846 | 0.93 |
|  | **b (Aβ40→ T1w/T2w-FLAIR)** | **Direct** | **0.0006** | **0.0002** | **0.004** |
|  | **c (VD → T1w/T2w-FLAIR)** | **Direct** | **0.057** | **0.025** | **0.023** |
|  | ab | Indirect | 0.0003 | 0.003 | 0.92 |
| **Model 2: VD Status → Aβ42→ T1w/T2w-FLAIR Ratio** | a (VD → Aβ42) | Direct | 0.094 | 0.307 | 0.31 |
|  | **b (Aβ42 → T1w/T2w-FLAIR)** | **Direct** | **0.009** | **0.004** | **0.019** |
|  | **c (VD → T1w/T2w-FLAIR)** | **Direct** | **0.057** | **0.025** | **0.025** |
|  | ab | Indirect | 0.001 | 0.003 | 0.75 |
| **Model 3: VD → Vascular Risk 3+ → T1w/T2w-FLAIR Ratio** | **a (VD → Vasc Risk 3+)** | **Direct** | **0.177** | **0.057** | **0.002** |
|  | **b (Vasc Risk 3+ → T1w/T2w-FLAIR)** | **Direct** | **0.056** | **0.017** | **0.001** |
|  | **c (VD → T1w/T2w-FLAIR)** | **Direct** | **0.053** | **0.026** | **0.041** |
|  | **ab** | **Indirect** | **0.01** | **0.004** | **0.023** |

Abbreviations: VD, vascular dementia; SE, standard error; Aβ, amyloid-beta.

Note: Model 1 evaluates whether the association between VD status and residualized T1w/T2w-FLAIR ratio is mediated through plasma Aβ40. Model 2 evaluates whether the association between VD status and residualized T1w/T2w-FLAIR ratio is mediated through plasma Aβ42. Model 3 evaluates whether the association between VD status and residualized T1w/T2w-FLAIR ratio is mediated through high vascular comorbidity burden (≥3 risk factors), additionally adjusting for plasma Aβ42. All mediation models were estimated using structural equation modeling (SEM) via the lavaan package and adjusted for age, sex, years of education, imaging site, cognitive diagnostic group, and days between MRI acquisition and clinical visit. Path a represents the effect of the predictor on the mediator; path b represents the effect of the mediator on residualized T1w/T2w-FLAIR ratio; path c represents the direct effect of the predictor on residualized T1w/T2w-FLAIR ratio after accounting for the mediator; ab represents the indirect (mediated) effect. Bold values indicate p < 0.05.

**Supplemental Table 13. Sensitivity Analysis: Primary Multivariable Model with ComBat-Harmonized T1w/T2w-FLAIR Residuals**

| **Term** | **B** | **Standard Error** | **P-value** | **CI_lower_95** | **CI_upper_95** | **B (Imputed)** | **P-value (Imputed)** |
| --- | --- | --- | --- | --- | --- | --- | --- |
| **Intercept** | **-0.37** | **0.074** | **<0.001** | **-0.52** | **-0.23** | **-0.34** | **<0.001** |
| **Age** | **0.0039** | **0.00096** | **<0.001** | **0.0020** | **0.0058** | **0.0033** | **<0.001** |
| Sex (Male) | 0.0058 | 0.016 | 0.71 | -0.025 | 0.036 | 0.011 | 0.43 |
| Education (years) | 0.0014 | 0.0016 | 0.39 | -0.0018 | 0.0047 | 0.00088 | 0.56 |
| Days Between Scan and Visit | -0.00048 | 0.00030 | 0.12 | -0.0011 | 0.00012 | -0.00034 | 0.21 |
| Site: Chonnam | 0.052 | 0.041 | 0.20 | -0.028 | 0.13 | 0.057 | 0.13 |
| Site: Gwangju | 0.00099 | 0.045 | 0.98 | -0.087 | 0.089 | -0.0054 | 0.89 |
| Site: Inha | 0.021 | 0.022 | 0.32 | -0.021 | 0.064 | 0.025 | 0.18 |
| Site: Pusan | -0.047 | 0.033 | 0.15 | -0.11 | 0.018 | -0.036 | 0.26 |
| Site: Samsung | 0.0075 | 0.023 | 0.75 | -0.039 | 0.054 | 0.031 | 0.14 |
| Site: Suwon | -0.022 | 0.020 | 0.26 | -0.060 | 0.017 | -0.010 | 0.58 |
| Group: MCI non-amnestic | 0.010 | 0.029 | 0.72 | -0.046 | 0.067 | 0.0090 | 0.73 |
| Group: MCI amnestic | 0.034 | 0.025 | 0.17 | -0.015 | 0.084 | 0.031 | 0.16 |
| **Group: AD** | **0.070** | **0.028** | **0.011** | **0.016** | **0.12** | **0.061** | **0.012** |
| **Group: VD** | **0.048** | **0.028** | **0.090** | **-0.0076** | **0.10** | **0.050** | **0.050** |
| Group: Other | 0.033 | 0.042 | 0.43 | -0.049 | 0.12 | 0.035 | 0.35 |
| **Plasma Aβ42** | **0.0078** | **0.0037** | **0.034** | **0.00058** | **0.015** | **0.0071** | **0.038** |
| Vascular Risk: 1 | 0.016 | 0.019 | 0.39 | -0.021 | 0.053 | 0.015 | 0.37 |
| Vascular Risk: 2 | 0.021 | 0.020 | 0.29 | -0.018 | 0.061 | 0.024 | 0.19 |
| **Vascular Risk: 3+** | **0.070** | **0.022** | **0.0017** | **0.026** | **0.11** | **0.068** | **0.00095** |

**Abbreviations:** AD, Alzheimer’s disease; Aβ42, amyloid-beta 42; CI, confidence interval; MCI, mild cognitive impairment; SCI, subjective cognitive impairment; VD, vascular dementia.

**Notes:** Sensitivity analysis re-fitting the primary Table 2 model after ComBat harmonization of global and white-matter T1w/T2w-FLAIR ratios, with site as the batch variable and age, sex, education, days between scan and visit, and clinical diagnosis preserved as biological covariates. T1w/T2w-FLAIR residuals were re-computed from the harmonized values prior to model fitting (residualization R² = 0.75). Reference categories: Group = SCI; Sex = Female; Site = Ajou. Multiple imputation by chained equations (MICE) was performed using the random forest method with m = 5 imputations. All other model specifications are identical to the primary analysis.

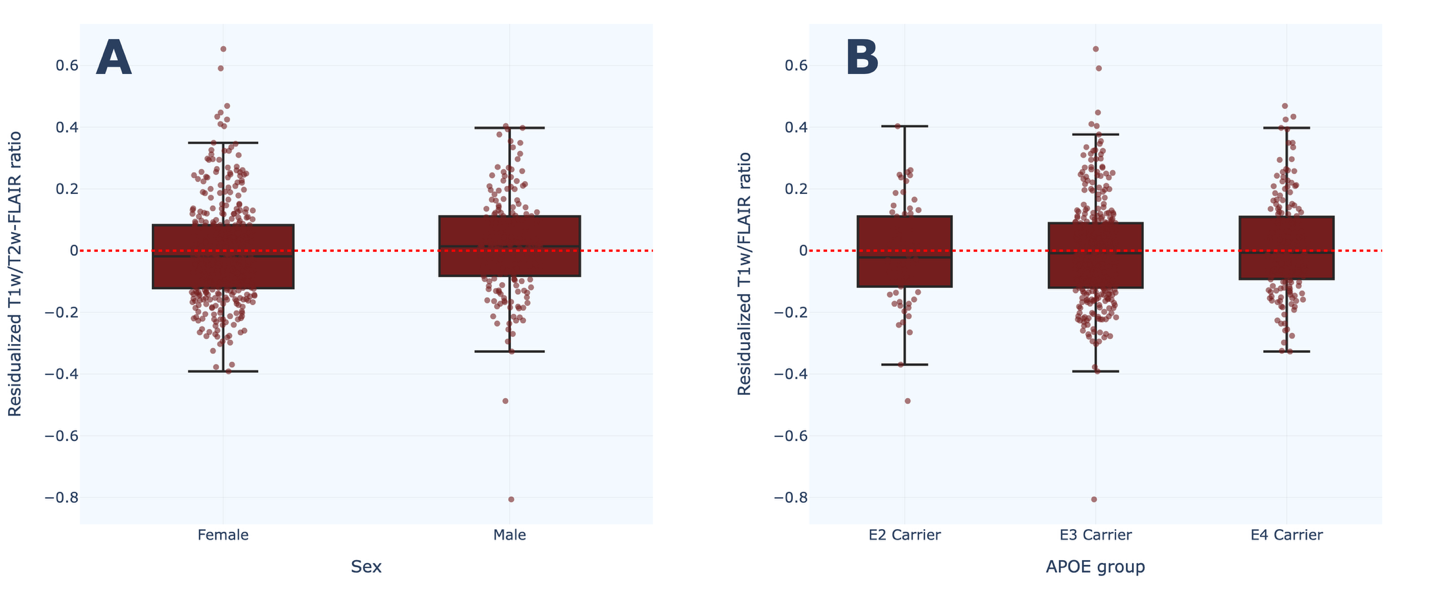
 **Supplemental** **Figure 1. Distribution of Residualized T1w/T2w-FLAIR Ratio by Sex and APOE Genotype**. Box plots showing the distribution of residualized T1w/T2w-FLAIR ratio across **(A)** sex and **(B)** APOE genotype groups. Individual data points are overlaid on box plots. The red dotted line at zero represents the expected T1w/T2w-FLAIR ratio after accounting for white matter contributions. Panel **A** demonstrates comparable distributions between females and males. Panel **B** shows similar T1w/T2w-FLAIR ratios across E2, E3, and E4 carriers. Box plots show the median (center line), interquartile range (box), and whiskers extending to 1.5× the interquartile range.

**Abbreviations:** APOE, apolipoprotein E
